# Methodological development of molecular endotype discovery from synovial fluid of individuals with knee osteoarthritis: the STEpUP OA Consortium

**DOI:** 10.1101/2023.08.14.23294059

**Authors:** Y. Deng, T.A. Perry, P. Hulley, R.A. Maciewicz, J. Mitchelmore, D. Perry, S. Larsson, S. Brachat, A. Struglics, C.T. Appleton, S. Kluzek, N. K. Arden, D. Felson, B. Marsden, B.D.M. Tom, L. Bondi, M. Kapoor, V. Batchelor, J. Mackay-Alderson, V. Kumar, L. S. Lohmander, T. J. Welting, D. A. Walsh, A.M. Valdes, the STEpUP OA Consortium, T. L. Vincent, F. E. Watt, L. Jostins-Dean

## Abstract

**Objectives:** To develop and validate a pipeline for quality controlled (QC) protein data for largescale analysis of synovial fluid (SF), using SomaLogic technology.

**Design:** Knee SF and associated clinical data were from partner cohorts. SF samples were centrifuged, supernatants stored at −80 °C, then analysed by SomaScan Discovery Plex V4.1 (>7000 SOMAmers/proteins).

**Setting:** An international consortium of 9 academic and 8 commercial partners (STEpUP OA).

**Participants:** 1746 SF samples from 1650 individuals comprising OA, joint injury, healthy controls and inflammatory arthritis controls, divided into discovery (n=1045) and replication (n=701) datasets.

**Primary and secondary outcome measures:** An optimised approach to standardisation was developed iteratively, monitoring reliability and precision (comparing coefficient of variation [%CV] of ‘pooled’ SF samples between plates and correlation with prior immunoassay for 9 analytes). Pre-defined technical confounders were adjusted for (by Limma) and batch correction was by ComBat. Poorly performing SOMAmers and samples were filtered. Variance in the data was determined by principal component (PC) analysis. Data were visualised by Uniform Manifold Approximation and Projection (UMAP).

**Results:** Optimal SF standardisation aligned with that used for plasma, but without median normalisation. There was good reliability (<20 %CV for >80% of SOMAmers in pooled samples) and overall good correlation with immunoassay. PC1 accounted for 48% of variance and strongly correlated with individual SOMAmer signal intensities (median correlation coefficient 0.70). These could be adjusted using an ‘intracellular protein score’. PC2 (7% variance) was attributable to processing batch and was batch-corrected by ComBat. Lesser effects were attributed to other technical confounders. Data visualisation by UMAP revealed clustering of injury and OA cases in overlapping but distinguishable areas of high-dimensional proteomic space.

**Conclusions:** We define a standardised approach for SF analysis using the SOMAscan platform and identify likely ‘intracellular’ protein as being a major driver of variance in the data.

**Strengths and limitations:**

- This is the largest number of individual synovial fluid samples analysed by a high content proteomic platform (SomaLogic technology)
- SomaScan offers reliable, precise relative SF data following standardisation for over 6000 proteins
- Significant variance in the data was driven by a protein signal which is likely intracellular in origin: it is not yet clear whether this is due to technical considerations, normal cell turnover or relevant pathological processes
- Adjusting for confounding factors might conceal the true structure of the data and reduce the ability to detect ‘molecular endotypes’ within disease groups

## INTRODUCTION

Osteoarthritis (OA) is a highly prevalent and disabling condition and arguably represents one of the greatest unmet clinical needs of all musculoskeletal conditions [1, 2], now recognised by the FDA as a ‘serious disease’ [3, 4]. OA is a disease of the synovial joints manifesting as localised, low-grade inflammation of the synovium, cartilage damage and subchondral bone remodeling [5], which lead to pain, stiffness and loss of function [6, 7]. Despite growing clinical demand and best efforts in pre-clinical models and translational studies to understand the underlying pathogenesis, target discovery and drug development for knee OA in humans have been slow [8]. Results from randomised clinical trials of putative disease-modifying osteoarthritis drugs (DMOADs) have been largely disappointi^[9^n, g^10]^ with a few treatments showing modest effects on cartilage preservation [11, 12]. OA might not be as in gledisease[13, 14] but rather a group of diseases with a similar clinical presentation but driven by distinct molecular pathways known as ‘endotypes’. These might determine the course of disease and in some cases predict response to treatment. It is presumed that molecular endotypes might relate to discernible patient characteristics and may help to explain the heterogeneity of OA ‘clinical phenotypes’ [15, 16]. Many cellular processes have been proposed as critical drivers in OA pathogenesis such as immune-mediated inflammation [17], mechanically-mediated inflammation (‘mechanoflammation’) [18], low/failed tissue repair [19, 20] and cellular senescence[21]. These in turn may relate to a broad range of aetiological factors that are associated with OA [22–26]. Efforts have been made to classify subgroups of people with OA based on epidemiological factors [27], with several clinically defined phenotypes now suggested in the literature[9, 28–30]. A recent systematic review of 24 studies reported that up to 84% of people with OA could be assigned to at least one of six phenotype^[2^s^8, 31]^. These clinical phenotypes are, however, not mutually exclusive, and are poor systematic classifiers because they are a mixture of overlapping demographic, clinical, radiographic, aetiological and systemic features. They currently have limited clinical applicability [29] and there is a paucity of data relating them to distinct molecular pathways or to clinical outcomes in OA[32].

Whilst there has been a plethora of studies of candidate molecules trying to identify diagnostic or prognostic biomarkers of OA, relatively few have used hypothesis-free approaches in large numbers of human biological samples to identify molecular endotypes [28, 33, 34]. Such collaborations are required to help move biomarker discovery forward [35]. Two broad matrices have been studied: blood (plasma or serum), which has the advantage of accessibility, and synovial fluid (SF), acquired by joint aspiration. SF has several advantages over blood for exploring molecular mechanisms.

Firstly, it has adjacency to joint tissues, and may reflect activities in synovium, bone as well as cartilage[36–38]. Secondly, concentrations of analytes within the SF provide an indication of biological activity and target tissue activation [39, 40]. Thirdly, the SF from a given joint is less confounded by disease at other sites than is, for example, blood. Finally, a number of analytes that are highly regulated in the SF are not reflected in the plasma [36, 41, 42].

The S ynovial fluid T o detect molecular E ndoty<u>p</u>es by U nbiased P roteomics in <u>OA</u> (STEpUP OA) Consortium was set up to address a primary objective: to determine whether there are detectable distinct molecular endotypes in knee OA, through a hypothesis-free, unsupervised proteomic analysis applying SomaLogic array technology [43] of SF from a large number of participants with, or at increased risk of, knee OA. SomaScan, an aptamer-based proteomics technology, offers the ability to measure large numbers of protein analytes from a small volume of biological fluid. However, detailed methodology is lacking for quality control (QC) and data analysis pipelines specifically tailored to SF.

SF presents analysis challenges due to its complex matrix which is rich in hyaluronan making the fluid viscous, variability in joint effusion volume between and within patients, and potential contamination with blood at time of aspiration. To combat some of these challenges, hyaluronidase treatment of the fluid post aspiration or lavage of the joint prior to aspiration have been utilised [42, 44].

In this study we describe the processing and analysis of SF, and the optimisation of a standardised quality control (QC) and analysis pipeline for these data. We evaluate performance of SF on the SomaLogic platform at scale for the first time and identify important technical confounders requiring adjustment prior to downstream analysis. Prespecified potential confounding factors included those relating to sample processing or to the sample itself, such as its age, number of freeze-thaws, visible blood staining and sample volume. These investigations were used to inform STEpUP OA’s primary data analysis plan (https://www.kennedy.ox.ac.uk/oacentre/stepup-oa/stepup-oa) and make our work replicable by others.

## METHODS

Details of consortium structure, governance and ethical approvals can be found in Supplementary Methods. Working groups oversaw key activities (Figure S1). Six participating sites with 17 participant collections (henceforth referred to as ‘cohorts’) including those with either knee OA or acute knee joint injury provided associated SF samples. Each had ethical approval (Table S1). In addition, the University of Oxford Medical Sciences Central University Research Ethics Committee (CUREC) granted ethical approval for the processing, storage and use of samples and linked data for this project on 1^st^ November 2019 (R67029/RE001).

### Participant eligibility criteria

All but one cohort had existing associated stored participant SF samples. Inclusion criteria were: i) evidence of a confirmed diagnosis of knee OA, or history of recent knee injury, ii) associated basic clinical information including (as a minimum) age at sampling, sex and indication of OA disease status, iii) a minimum volume of SF (90 µl, ideally 200 µl) and iv) SF had been centrifuged between 1800-3000g, prior to supernatant storage at −80 °C. Exclusion criteria were: i) additional forms of arthritis e.g. gout, rheumatoid arthritis, psoriatic arthritis, as determined by host investigator; ii) confounding medical conditions e.g. concurrent infection, cancer; iii) confounding treatments e.g. index knee surgery in the preceding 6 months, index knee steroid injection in preceding 3 months; iv) chemotherapy and; v) significant deviation in storage procedure (e.g. freezer drop-out defined by host investigator).

### Sample processing and SomaLogic assay

Consortium samples: 1746 SF samples were eligible and processed for STEpUP OA. A STEpUP participant ID number (PIN) and related unique sample identification number (SIN) were generated for each participant and their associated sample(s). Sample processing was performed in Oxford in four tranches over a 24-month period. For analysis by SomaLogic, SF enzymatic digestion, using hyaluronidase, was carried out (Supplementary methods). Briefly, sufficient bovine testicular hyaluronidase (4mg/ml; Sigma-Aldrich) for the entire project was reconstituted as a single batch from a single lot number and frozen in aliquots until use. For each tranche, batch processing was performed over consecutive working days, i.e. over as short a time as possible. Briefly, a batch of SFs was thawed, centrifuged at 3000g at 20°C for 25 minutes and 175 µl of SF supernatant diluted 1:2 with the same volume of hyaluronidase solution and agitated at room temperature for 1 hour, followed by further centrifugation for 5 minutes [42]. Supernatants were aliquoted and stored at −80°C and transferred on dry ice by temperature-controlled shipping to SomaLogic (single shipment per tranche).

Consortium controls/QC samples: Equal volumes of SF samples from 6 participants per group were used to generate single batches of hyaluronidase-treated ‘pooled samples’ for each of OA and knee injury at the start of project. Subaliquots of these then acted as internal QC controls, being run on each SomaLogic plate, enabling calculation of intra-assay and inter-assay coefficients of variation (CVs), as well as assessing effects of freeze thaw (a multiple freeze-thawed aliquot), frozen storage of hyaluronidase (an untreated aliquot freshly treated with frozen hyaluronidase during processing of each tranche) and centrifugation (an additional unspun pooled sample, from 6 unspun OA SF samples). A further 18 samples (split at the time of collection, with the paired aliquot remaining ‘unspun’) were included to further examine the effects of centrifugation. 42 other ‘comparator’ samples were included (disease-free controls from non-painful knees or from normal joints at amputation/post-mortem; samples from individuals with definite inflammatory arthritis). Three samples from three separate participants were re-processed under 3 different temperature conditions and re-analysed to examine the effects of laboratory re-processing. A subgroup of the freshly collected samples were processed specifically to test generalizability to OA SF which had not been centrifuged (‘unspun’) (n=18). 235 unspun OA samples were subsequently included in the replication analysis.

Samples were assayed on the SomaLogic SomaScan Discovery Plex V4.1 by SomaLogic, in Boulder, US. All samples from all 4 tranches were processed as a single batch on twenty-two sequential 96-well plates in January 2022. All samples were randomised within and between plates whilst ensuring appropriate controls on each plate. Each plate included 83 participant SF single samples; one pooled OA sample; one pooled knee injury sample; five plasma calibrator samples; three plasma QC samples and three blanks per plate. The SomaScan platform quantified 7,596 synthetic DNA SOMAmers (for 7289 human targets) (Slow Off-rate Modified Aptamers [45, 46]) that bound to 6596 unique human proteins. The generated SomaScan protein quantification was securely transferred from SomaLogic to Oxford as .adat files.

Additional sample metadata, both from collecting sites (where available) and those generated in Oxford and at SomaLogic, included: sample blood staining (by visual staining defined by host investigator at time of collection); initial centrifugation of the sample; number of previous freeze thaws; the date of laboratory processing; batch and order of processing; the plate and position of the sample. These sample metadata were defined as technical confounders in our QC pipeline (Table S2).

### Clinical data

Pseudonymised associated participant clinical data were transferred from participating sites to Oxford, linked to their consortium PIN, mapped to variables where necessary and uploaded to a REDCap database (Research Electronic Data Capture, Vanderbilt University, US) [47], hosted by the University of Oxford. Data integrity and completeness were ensured using a data dictionary, data entry constraints and a combination of automated, systematic and random checks by two of the study team.

A consortium working group oversaw all aspects of data management including definition of variables and associated data dictionary, data harmonisation and design of the database (Figure S1).

Informed by their relative clinical importance, by data availability and by iterative review, a core clinical dataset (a subset of the data dictionary) was defined: the first phenotype release “Pheno 1” (demographic data and harmonised measures of radiographic disease severity) and the second release “Pheno 2” (dichotomous and continuous harmonised patient-reported outcome measures for knee pain[48]). (Table S2 & Supplementary methods).

### Data QC approach

Methods to develop QC and data analysis pipelines prior to the primary discovery analysis were pre-defined in the Quality Assurance plan (https://www.kennedy.ox.ac.uk/oacentre/stepup-oa/stepup-oa). This QC pipeline aimed to validate methods for standardisation of the data, through a series of normalisation steps (given that SF was a non-standard matrix on SomaScan), correction for technical confounders and filtering based on pre-defined quality thresholds for SOMAmers, proteins and samples. The approach included pre-defined data exploration, though where issues were found, these were iteratively investigated and findings used to refine the QC pipeline. This approach was informed by our prior published work [42], SomaLogic expertise, initial consortium pilot work on 435 samples previously assessed on an earlier version (4.0) of the SomaScan platform, and subsequent QC work within this dataset.

The usual SomaScan analysis pipeline for plasma involves a series of standardisation procedures to reduce nuissance variance, using plasma calibrator and plasma QC samples included on plates to reduce the effect of technical factors across samples and plates [49]. This routine standardisation of the SomaScan relative fluorescence units (RFU), adjusted by the protein’s dilution factor used in the SomaScan assay (the “dilution bin”), was applied in a stepwise way, using i) hybridisation control normalisation (to remove well-to-well variation due to different rates of hybridisation between SOMAmers and fluorescence probes using spiked-in control SOMAmers); ii) plate scaling using plasma calibrators (to remove variation in overall intensity between plate normalisation (to decrease variation due to total fluorescence intensity between samples); and iv) calibration (using plasma calibrator samples of known concentration to rescale each protein and reduce assay differences between runs). Each normalisation step was tested in a sequential manner.

After optimised standardisation, the R package*limma* was used to adjust proteins for a number of pre-specified continuous covariates, as described below. The batch correction method ComBat (in sva R package) was applied [50] to adjust the mean and variance of each protein for batch effects in datasets where the batch covariate, such as plate, is known [44] (again, as described below).

Based on our initial QC assessments, to quantify and adjust for differences in the contribution of intracellular proteins to the proteome, we defined an Intracellular Protein Score (IPS) for each sample *i*, as a weighted sum of protein concentrations using the equation

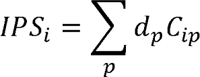

where d_p_ is the Cohen’s d for the difference in concentration for protein P between paired spun and unspun samples, and c_ip_ is the log concentration of protein *P* in sample *i*.

Checks of assay performance and biological validity were carried out by measuring repeatability using the pooled samples on each plate, use of metadata for prespecified technical confounders, and comparison with previously generated quantitative immunoassay data (R&D or Meso Scale Discovery, available for 60 OA and injury SFs amples (without hyaluron idasetreatment) for 9 overlapping proteins (Table S3).

### Statistics and Analysis

Principal Components Analysis (PCA) was used to visualize proteome-wide patterns of variation in the data, with further visualisation of Principal Components (PCs) with Uniform Manifold Approximation and Projection (UMAP) 2-dimensional plots (UMAP applied to the set of top PCs that explained >80% of total variation). Various other bioinformatic, descriptive and statistical techniques were employed to test the quality of the data. We checked:

- Inter-assay repeatability – the %CV of each protein, i.e. the ratio of the standard deviation of the concentration to the mean of the concentration within repeated samples, and the proportion of non-technical variation (R ^2^) for each protein, estimated as one minus the square of the ratio of the variance in repeated samples to the variance in non-repeated SF samples.
- The effect of freeze-thawing on normalised RFU signal (measured by %CV between repeatedly freeze-thawed and non-freeze-thawed samples).
- The effect of centrifugation on normalised RFU signal (by estimates of correlation between, and differential abundance of, proteins in unspun and spun samples, using correlation tests and paired t-tests respectively).
- Assay accuracy – comparing SomaScan normalised RFU signal with existing quantitative immunoassay data (by Pearson correlation coefficients).
- Effects of each technical confounder on standardised RFU signal (from combined cohorts). Linear regression analyses were applied to identify the most significant principal components (PCs) and proteins associated with technical confounders (Bonferonni adjusted p<0.05).

In addition, SOMAmers and samples of insufficient quality were removed as follows:

*SOMAmer filtering:*

- SOMAmers which were highly associated with pre-specified confounders (Bonferroni adjusted p<0.05) were removed (Table S4).
- SOMAmers from non-human organisms or control SOMAmers (including Spuriomer, hybridisation control elution, deprecated, non-biotin, and non-cleavable) were excluded.
- The estimated proportion of non-technical (i.e. biological) variation R ^2^ was calculated using pooled SF samples (OA, knee injury), defined as (1 – V _Pooled_/V_total_)^2^, where V _Pooled_ and V_total_ are the variances (V) in pooled and non-pooled (including all individual) samples respectively. If

R^2^ for a given SOMAmer accounted for less than 50% of total variation in either OA or knee injury it was removed.

*Sample filtering:*

- If a sample had more than 25% of protein values above or below the upper or lower limits of detection respectively, the sample was removed.
- (This was applying lower and upper limits of detection (LOD) (defined by SomaLogic), where lower LOD was: *[median concentration of blanks] + 4.9 x [median absolute deviation of blanks]*, based on three blanks (i.e. buffer only) per plate. Upper LOD was defined as 80,000 RFU).
- Identification of outliers by PCA: samples that were beyond 5 standard deviations (SDs) from the center of the principal component space (made up of PCs that explained at least 80% of variation) were removed.
- Identification of total signal intensity outliers: samples that were beyond 5 SDs from the mean in total RFU distribution were removed. Total RFU of per sample was defined as the sum of all the RFU values of that sample.
- Samples that were flagged by SomaLogic’s in-house QC process were removed.

Pearson correlation coefficients (with 95% confidence intervals) are given for correlations and all analyses were carried out in R (version 4.3.1), unless otherwise stated.

## RESULTS

Of 1746 unique participant samples included, tranches 1&2 were designated the Discovery analysis dataset (comprising 1045 samples), and tranches 3&4 the Replication analysis dataset (701 samples).

### Selection of Each Normalisation Procedure

To define the optimal QC pipeline, selection of appropriate standardisation procedures was needed. These had been previously optimised by SomaLogic for plasma samples. We set out to test how well these procedures performed in all 1746 SF samples with 7596 SOMAmer features.

We measured the impact of technical variation in each normalisation step (see methods) on assay performance in a sequential manner, firstly by measuring effects on the mean %CV and the mean R ^2^ across all proteins for the pooled sample replicates across all plates (Figure 1A&B). Comparing each to the raw RFU data, %CVs ranged from 11.65% to 16.49% for pooled OA samples (n=22) and 10.56% to 16.63% for pooled injury samples (n=22). Mean R ^2^ ranged from 77.70% to 88.98% and 82.59% to 89.10% for pooled OA and injury samples respectively, suggesting a high level of repeatability. A decreasing trend in mean %CVs suggested that the routine normalisation steps improved measure repeatability with the exception of median normalisation (Figure 1A&B). Removing median normalisation from the standardisation procedure resulted in a mean %CV of 13.6% and a mean R^2^ of 88.98% for pooled OA samples and %CV of 14.34% and a mean R^2^ of 89.1% for pooled injury samples (Figure 1A&B).

**Figure 1.**
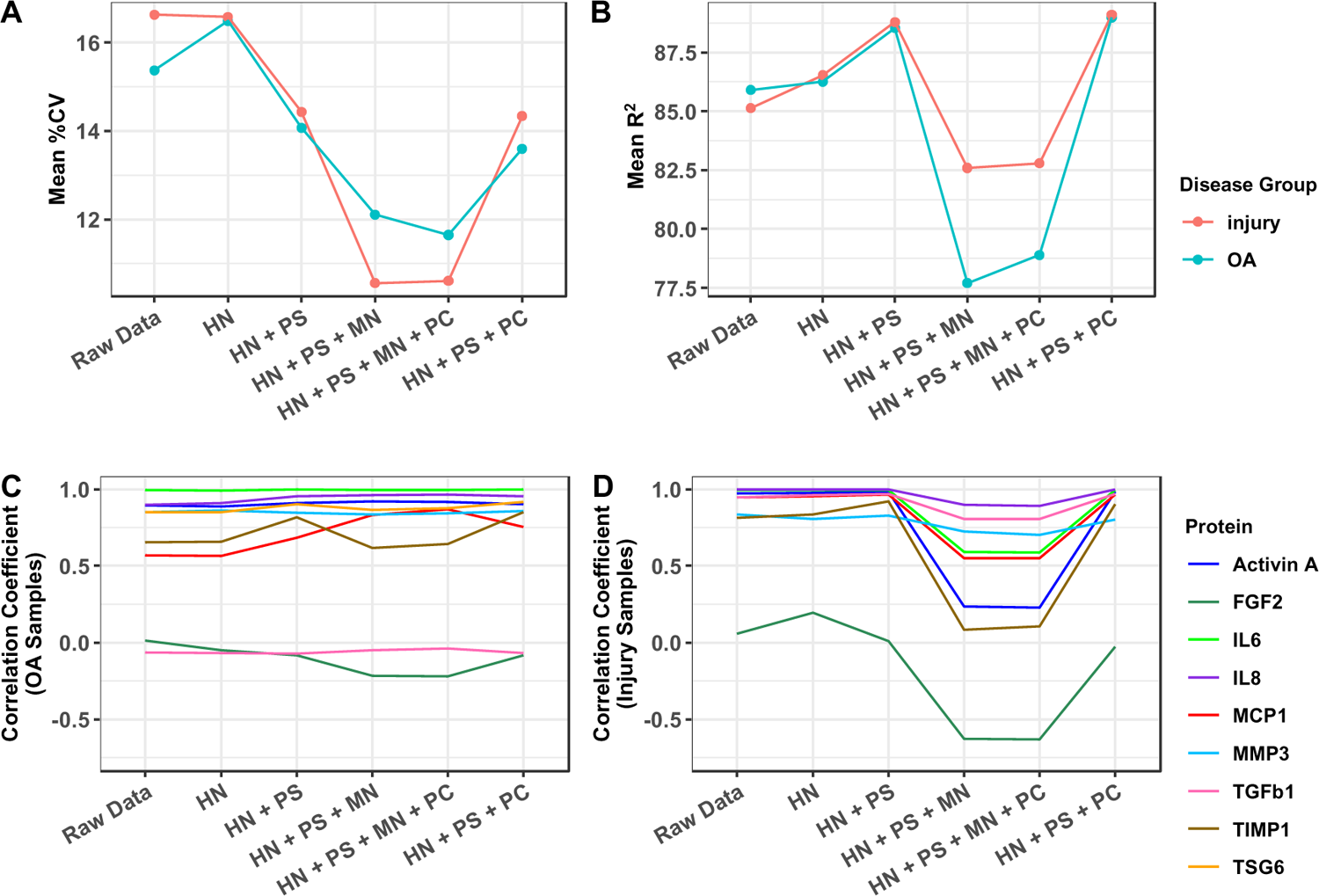
Assessment of the effects of each standardisation step on (A) mean %CV and (B) mean R^2^ across all proteins for pooled sample replicates, stratified by OA and acute knee injury respectively. Assessment of Pearson correlation coefficients between protein expression in samples measured by the SOMAscan platform and by prior immunoassay for nine select proteins across normalisation steps for (C) OA and (D) acute knee injury. The normalisation steps included hybridisation normalisation (HN), plate scaling using plasma calibrators (PS), median signal normalisation (MN) and calibration using plasma calibrators (PC). Correlation between the RFUs (SOMAscan) and absolute concentrations for the nine proteins across the two methods are shown. RFUs, relative fluorescence units; %CV, % coefficient of variation; osteoarthritis, OA; Activin A, Inhibin beta A chain; FGF2, Fibroblast growth factor 2; IL6, Interleukin-6; IL8, Interleukin-8; MCP1, C-C motif chemokine 2; MMP3, Stromelysin-1; TGF β1, Transforming growth factor beta-1; TIMP1, Tissue inhibitor of metalloproteinase 1; TSG6, Tumor necrosis factor-inducible gene 6.

Correlation of the SomaScan assay data with nine selected analytes measured in the same SF sample by immunoassay was generally high for most analytes (Figure 1C&D). However, upon normalisation steps, we saw a similar effect, with median signal normalisation reducing the correlation with these validation measurements, particularly in the injury samples (Figure 1D). Based on these combined data, we chose to use SomaLogic’s existing standardisation procedures, though omitting median normalisation from our SF standardisation pipeline (i.e. employing hybridisation normalisation, plate scaling, and plate calibration using SomaLogic’s plasma calibrators).

### Identification and correction of confounding factors

After conducting standardisation, principal component (PC) analysis identified one dominant component (PC1) that explained 48% of variation in the data (Figure 2A). This principal component was positively correlated with almost all proteins measured, with a median correlation coefficient of 0.70, with the highest correlations seen with low abundance proteins (Figure 2B). We were able to rule out a total protein effect, due to a low correlation (−0.039) with standard high-abundance markers such as albumin concentration. Examining the proteins that drove this signal, we found that lower protein abundance was the strongest independent predictor of correlation (Table S5) with PC1 (p*<* 2.23e-308), with the next most significant predictors being whether proteins were predicted not to be secreted (p=3.98e-10) and proteins that were identified as nuclear and not secreted (p=1.64e-9). This led to the hypothesis that PC1 was capturing an effect of intracellular proteins, perhaps reflecting cell turnover or due to the presence of microvesicles. A strong intracellular signal was confirmed by showing that PC1 was consistently reduced in spun samples when comparing paired SF samples (from the same parent SF, n =18) that had been split into two and either spun or left unspun immediately after joint aspiration (Figure 2C).

**Figure 2.**
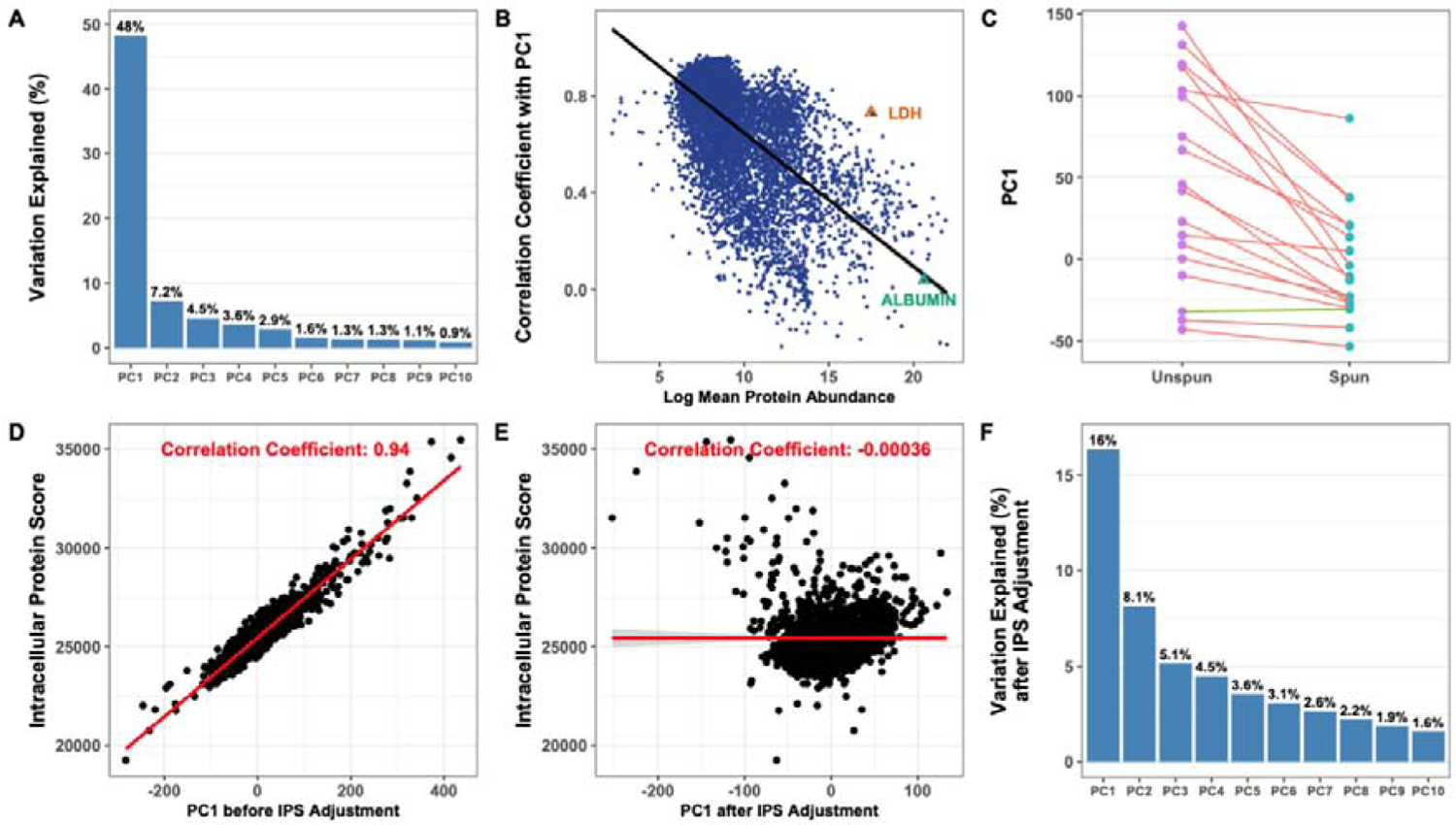
(A) Variation explained (%) by the top 10 PCs derived from the standardised log abundance proteomic data. (B) Correlation between PC1 and protein abundance, with two high-abundance proteins (albumin, a soluble serum protein, and LDH, an intracellular protein) marked. Protein abundance is calculated as the standardized RFU for each protein adjusted by the protein’s dilution factor used in the SomaScan assay (the “dilution bin”). (C) Comparison of variation explained (%) by PC1 between 18 pairs of SF samples that were centrifuged (spun) or not (unspun) after aspiration and prior to freezing, with paired samples from the same participant joined by separate lines. Red lines show samples that had an increased PC1 prior to spinning, and the green line where it was decreased. Correlation between PC1 and intracellular protein score (D) before and (E) after IPS adjustment. (F) Variation explained by the top 10 PCs derived from the batch corrected and IPS adjusted log abundance proteomic data. In all cases, correlation is measured using the Pearson correlation coefficient. IPS, Intracellular Protein Score; PC, principal component; LDH, Lactate dehydrogenase.

To quantify the contribution of intracellular proteins, we derived an Intracellular Protein Score (IPS) as the weighted sum of relative protein concentrations. For weights, we calculated a Cohen’s d from the 18 paired spun and unspun samples. This score correlated very highly with PC1 (Figure 2D). We used this score in a linear regression model adjusting for the contribution of intracellular proteins, which removed the correlation between IPS and PC1 (Figure 2E), reduced the variance explained by PC1 to 16% (Figure 2F), and removed the correlation of “non-secreted nuclear protein” with PC1 (Table S5).

This intracellular contribution to the SF proteome did not correlate strongly with any of our pre-defined technical confounders. However, it explained a large proportion of variation in our data, which ran the risk of swamping more subtle protein signatures or molecular endotypes, if present. We thus decided to include, in addition to the standardised data set without IPS adjustment, a co-primary dataset, applying IPS adjustment to each protein as part of our STEpUP OA Data Analysis Plan (https://www.kennedy.ox.ac.uk/oacentre/stepup-oa/stepup-oa).

We also found a strong ‘bimodal’ signal on PC2 of the data (Figure 3A&B) whereby a large number of SOMAmers (N=4030 at Benjamini–Hochberg (BH) adjusted p<0.05) were present at either very low or very high relative signal in a given sample. Further investigation showed that PC2 was highly correlated with the technical variable ‘laboratory processing batch’ (p<2.2E-308). Investigation of exemplar proteins displaying this behaviour showed that the bimodal signal followed sample processing order, usually (but not always) between laboratory processing batches (Figure 3C). The effect became stronger over time (Figure 3C). Re-analysing (at SomaLogic) previously laboratory processed (hyaluronidase treated) samples gave the same result (data not shown). However, when new aliquots of three sequential samples which had differing bimodal status were reprocessed by the Oxford laboratory and re-analysed, all three reprocessed results had a shared bimodal status.

**Figure 3.**
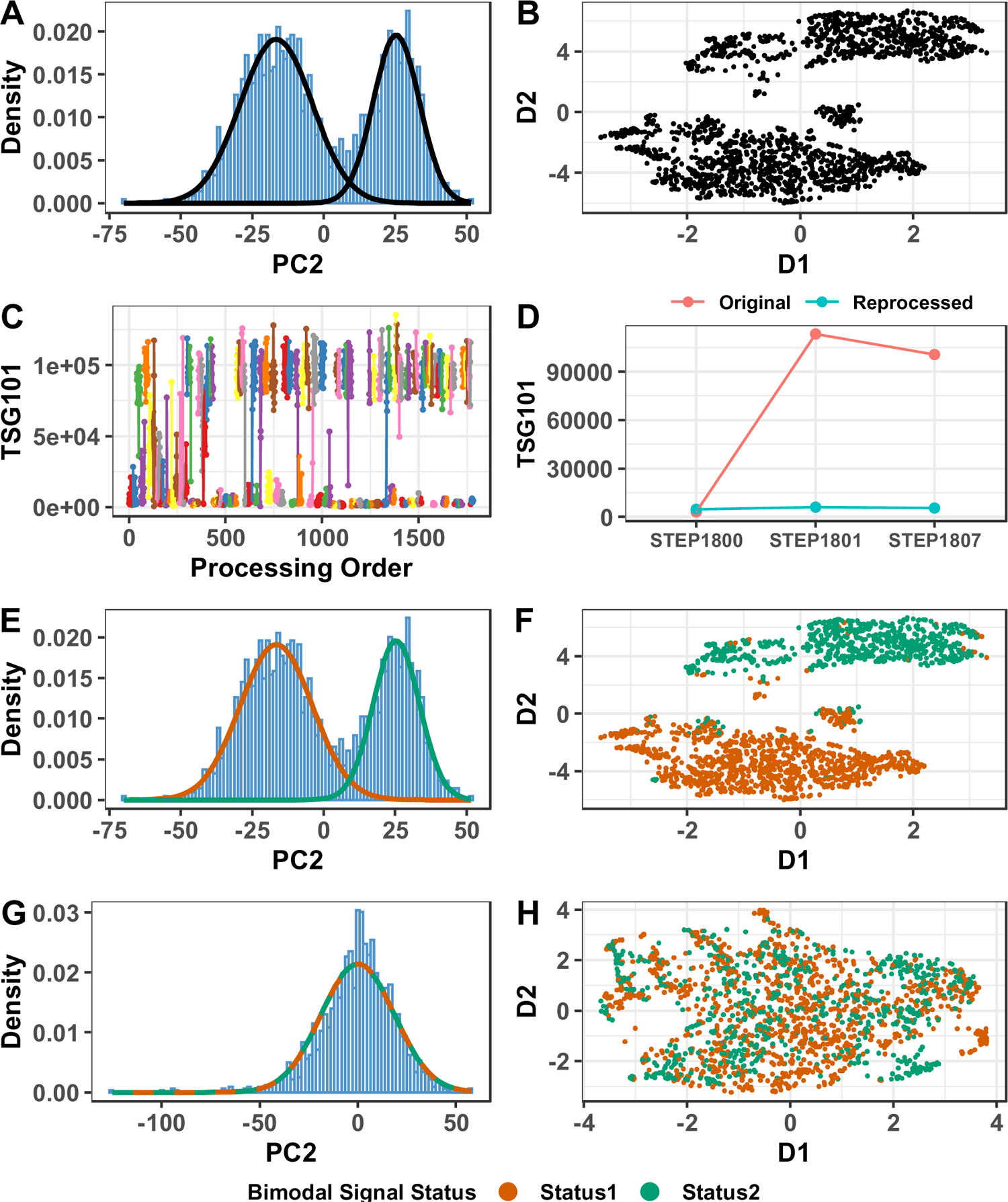
(A) Distribution of the second principal component (PC2) derived from the standardised log abundance data, showing a bimodal distribution. (B) UMAP visualisation of two reduced dimensions (D1 and D2) of the top PCs of the standardised log abundance data. (C) Example of a strongly bimodal protein measurement, TSG101, RFU (y-axis) against Oxford laboratory processing order (x-axis) and coloured by laboratory processing batch (with only points within the same processing batch connected by lines). Note that the ‘flipping’ between high and low signal status occurred primarily when processing batch changed, and only rarely within processing batch. This effect was particularly strong among sample batches that were processed later in processing order. (D) The same example protein measurement for three independent SF samples before (original) and after they were re-processed and re-assayed, showing that bimodal status changed after laboratory re-processing. (E) Distribution of PC2 derived from standardised log abundance data, showing the two probability density functions of the Gaussian Mixture Model used to classify samples into the two bimodal signal status groups. (F) UMAP visualisation of two reduced dimensions (D1 and D2) of the top PCs of the standardised log abundance data, colored by the inferred bimodal signal status. (G) Histogram of PC2 of the batch corrected log abundance data, with the now near-identical distributions of the two bimodal signal status groups shown as colored lines, (H) UMAP visualisation on two reduced dimensions (D1 and D2) of the top PCs of the batch corrected log abundance data, colored by the inferred bimodal signal status. RFUs, relative fluorescence units; PC, Principal Component; TSG101, Tumor susceptibility gene 101 protein; UMAP, Uniform Manifold Approximation and Projection.

This indicated that this was due, in some way, to our laboratory sample processing (hyaluronidase treatment) (Figure 3D). We hypothesised this might be due to sample temperature differences prior to hyaluronidase treatment, but further experiments did not corroborate this (data not shown).

Neither was this thought to be due to the stability of frozen hyaluronidase enzyme as the pooled OA sample, freshly processed during each tranche with frozen stored hyaluronidase, showed little variability over time (Figure S2).

We applied a Gaussian Mixture Model to PC2 to classify samples into high or low protein status, reflecting their bimodal signal (Figure 3E), which produced visually plausible assignments on PCA and UMAP (Figure 3E&F). To attempt to reduce this undesired variance, we carried out batch correction by samples’ PC2 bimodal signal status using the ComBat method[50].This correction reduced the impact of the bimodal signal considerably (Figure 3G&H) and was adopted into our QC pipeline.

We also discovered a significant influence of plate on a number of proteins (n=1927) at BH adjusted p<0.05). Our samples were randomised to plate, so this was unlikely to cause significant confounding in downstream analyses, but to reduce technical variation we also applied batch correction for plate by ComBat at the same time as correcting for the bimodal signal.

We assessed the impact of these adjustments described above using the immunoassay comparison data. While the IPS-adjustment reduced the dominance of PC1, it also had a negative impact on the correlation coefficients between SomaScan and prior immunoassay results of the select analytes (Figure 4). This was particularly evident in the injury group, where the correlation between the two measures for 4 out of 9 proteins (IL6, IL8, TGFß1, TIMP1) changed from strongly correlated to weakly or not correlated (Figure 4B). Interestingly, some of the measured cytokines (which had been selected because of their putative disease relevance) such as MCP1, IL-8 and TGFß1, correlated with the intracellular protein score (Table S6). Batch correction for plate and bimodal signal status (as part of our optimised standardisation) was seen to have minimal impact on immunoassay agreement (Figure 4).

**Figure 4.**
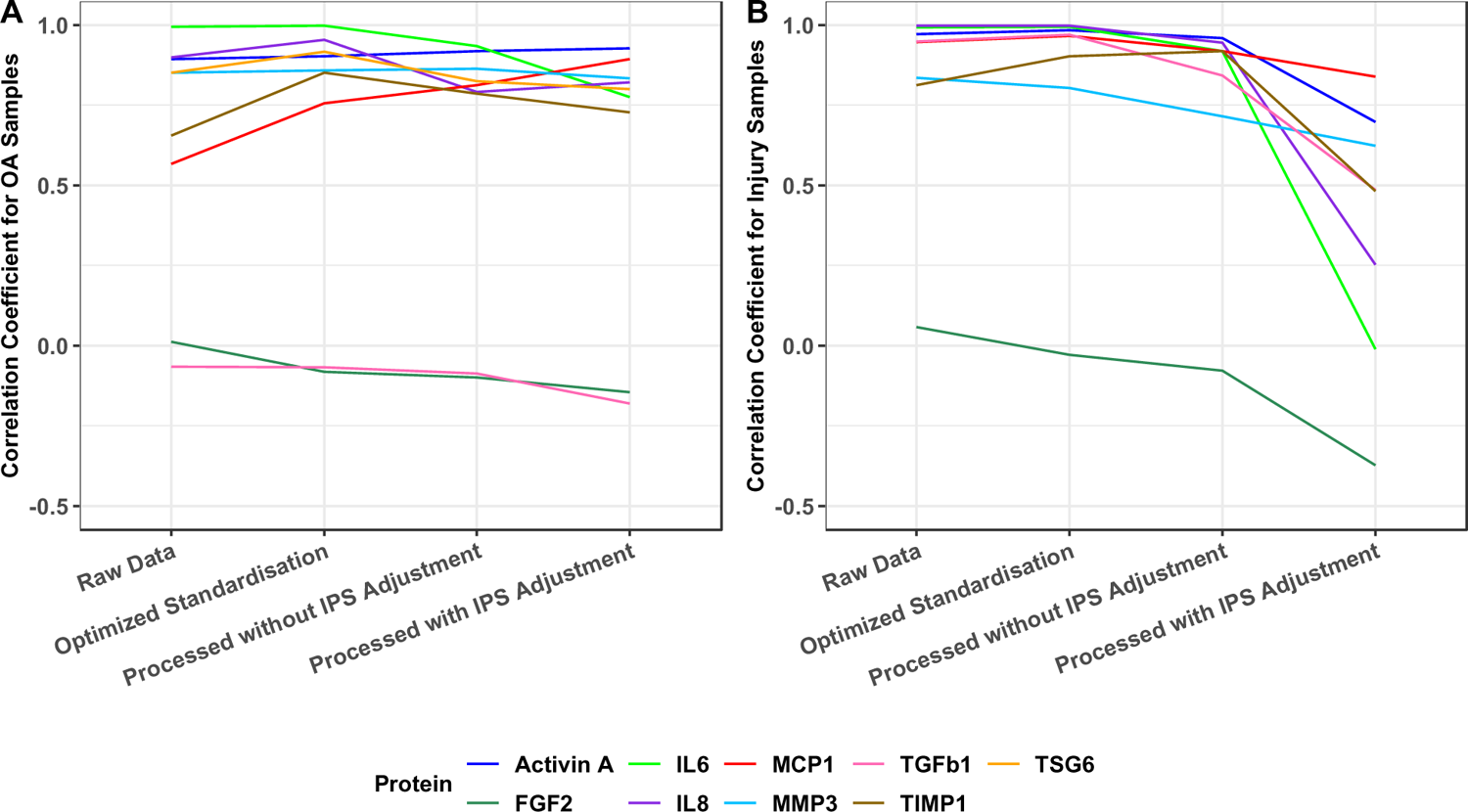
Correlation between SOMAscan relative frequency abundance (RFU) and abundance measured using orthogonal immunoassays for 9 selected proteins at different stages of SOMAscan data processing, for (A) osteoarthritis and (B) acute knee injury samples. Correlation was measured using the Pearson correlation coefficient. Raw data refers to the raw RFUs without any processing, optimised standardisation was the data standardised using our selected optimal normalization steps (Figure 1), processed without IPS adjustment refers to data that has been batch corrected for bimodal signal status and plate but not IPS adjusted, and processed with IPS adjustment refers to samples that have undergone both batch correction and IPS adjustment. IPS, Intracellular Protein Score; Protein name abbreviations as in Figure 1.

### Description and reduction of pre-defined technical confounding by protein filtering

In addition to identifying badly performing SOMAmers and samples for filtering according to assay performance (see methods), we also identified filters based on pre-defined technical confounders (Table S2).

Confounding technical factors were dealt with in different ways. Samples showed systematic biases in signal intensity plate position (Table S7, ‘Plate position’), but this was also deemed unlikely to confound downstream analyses because samples were randomised across and within plates, so its effect was not adjusted for. Blood staining (Table S7, ‘Visual blood staining’) and sample volume were both drivers of IPS, but we felt that both could contain biological signals of relevance, so they were not adjusted during QC but were considered covariates in the downstream analyses. Sample age (Table S7, ‘Sample age’) could introduce technical variation, therefore significantly associated proteins were removed by filtering (Table S4). Freeze-thawing was also shown to be a potential technical confounder (Table S7, ‘Sample freeze thaw cycles’). This was investigated further.

For freeze-thawing, we had repeatedly freeze-thawed (five times per sample) one aliquot each of the pooled OA and the pooled injury samples. This had an effect, particularly in the injury samples. However, the majority (77%) of proteins retained a good %CV (<20%) even after five freeze-thaws (Figure S2), suggesting that such samples remained usable. Technical variation brought about by freeze-thaw was nonetheless adjusted for by filtering out significantly associated proteins following Bonferroni correction (Table S4).

### Assessment of centrifugation effect on protein measurements

Although most of the samples had been centrifuged prior to initial storage as per our eligibility criteria, 240 samples included in the replication analysis were unspun. In anticipation of this we assessed further the impact of centrifugation on the 18 pairs of samples that had either been spun or left unspun at time of collection. We compared the SomaScan data to identify proteins that changed upon centrifugation (Figure S3). The effect of centrifugation on the data depended on whether the data were adjusted for IPS. The unadjusted data showed that centrifugation status was a major driver of variation across the paired samples, with a significant correlation with PC1 (Figure S3A, paired t-test p=0.0066), but after adjusting for IPS, the top PCs were no longer driven by spun status (Figure S3B, paired t-test p=0.2089). Centrifugation impacted the concentration of a large number of individual proteins in both IPS-unadjusted (n=5638, 74%, at BH adjusted p<0.05) and, to a lesser extent, IPS-adjusted data (n=3731, 49%, at BH adjusted p<0.05), although the majority of proteins were significantly correlated between the paired spun and unspun samples (n=6402, 85% in unadjusted data and n=4558, 60% in IPS adjusted data, Figure S3C&D respectively).

We concluded that spun and unspun samples were comparable (in that they captured similar information), but that any analysis that included both types together would need to adjust for systematic shifts in abundance and the small numbers of uncorrelated proteins. In our discovery and replication analysis plans relating to our primary analysis, only spun samples are therefore considered, with unspun samples used for secondary sensitivity analyses.

### Effect of blood staining on protein measurements

A subset of samples had information on blood staining, graded by visual inspection at the time of joint aspiration, prior to centrifugation. As shown in Table S7, the presence of blood measured in this way was a significant driver of protein variation. It was also a potential biological driver as haemarthrosis is common after significant joint injury and is known to be pro-inflammatory and associated with persisting knee symptoms [51–53]. Visual blood staining could reflect presence of either intact or lysed red blood cells. The analyte haemoglobin A (HBA) correlated reasonably well with visual blood staining grade prior to adjustment for IPS (Figure S4A), and less so after adjustment for IPS (Figure S4B). The log concentration of HBA relative abundance level (without IPS adjustment) was subsequently used as a measure of blood content, as a covariate in downstream analyses.

### Validation of data quality after QC

Following application of filters, 1720 samples and 6290 SOMAmers (features) remained. The total numbers of samples and proteins filtered out are shown in Table S4. After filtering, median %CV of pooled OA and injury samples remained relatively unchanged at 11.25 and 12.42 respectively (Figure S5). An overview of the end-to-end data processing and quality control pipeline, from raw data to final filtered data, is shown in Figure 5.

**Figure 5.**
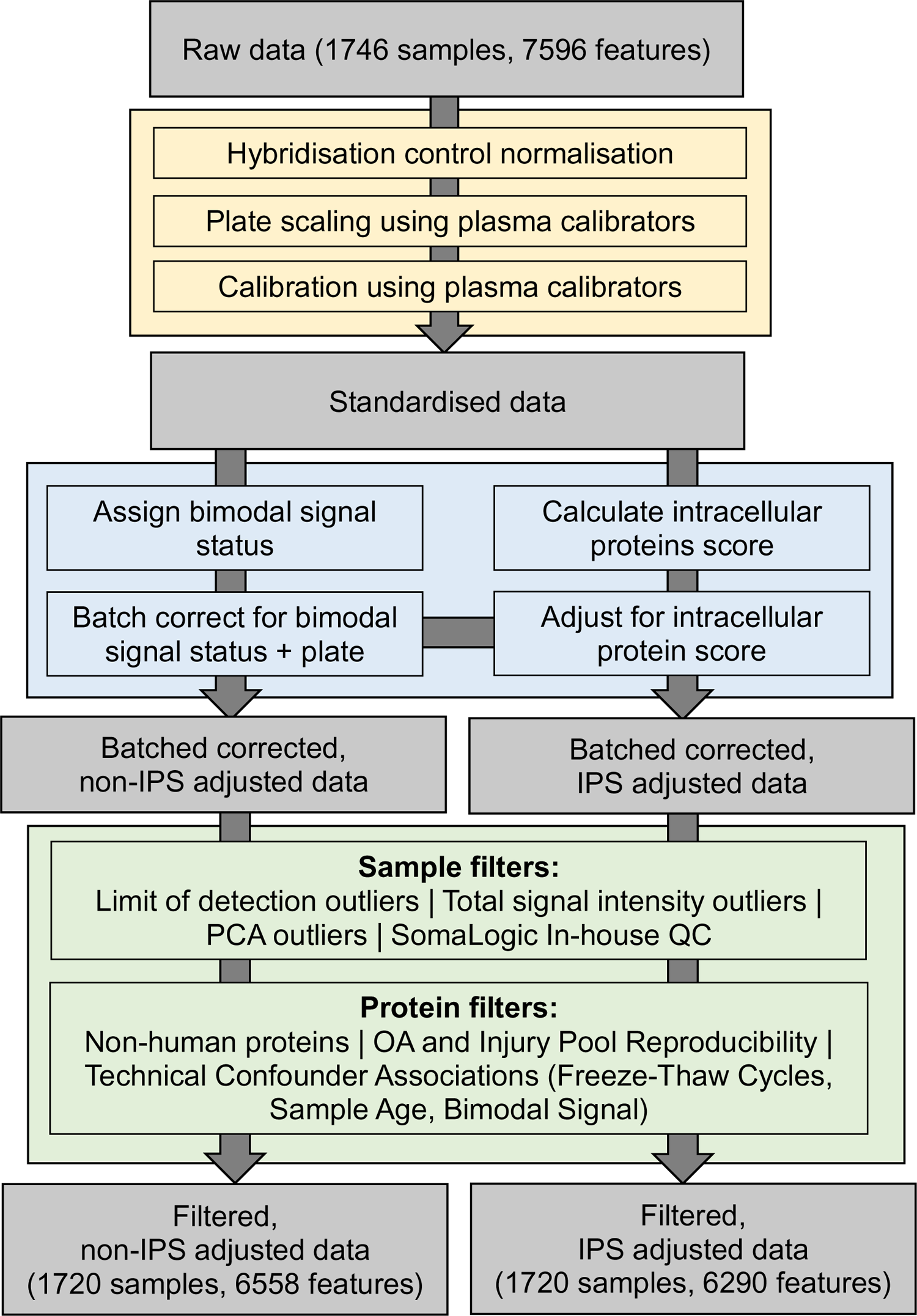
Overview of the final data processing and quality control pipeline for synovial fluid SOMAscan data used by the STEpUP OA consortium, broken down into three stages: standardisation (yellow box), technical confounder correction (blue box) and filtering (green box). More details on filtering thresholds, and the number removed by each filter, can be found in Supplementary Table S4.

The association of all these variables with the top 10 PCs of the standardised, bimodal signal corrected data after filtering is shown in Table S7. The strongest associations in the IPS adjusted, filtered data are shown in Figure 6. IPS adjusted non-filtered, and non-IPS adjusted data are shown in Figure S6.

**Figure 6.**
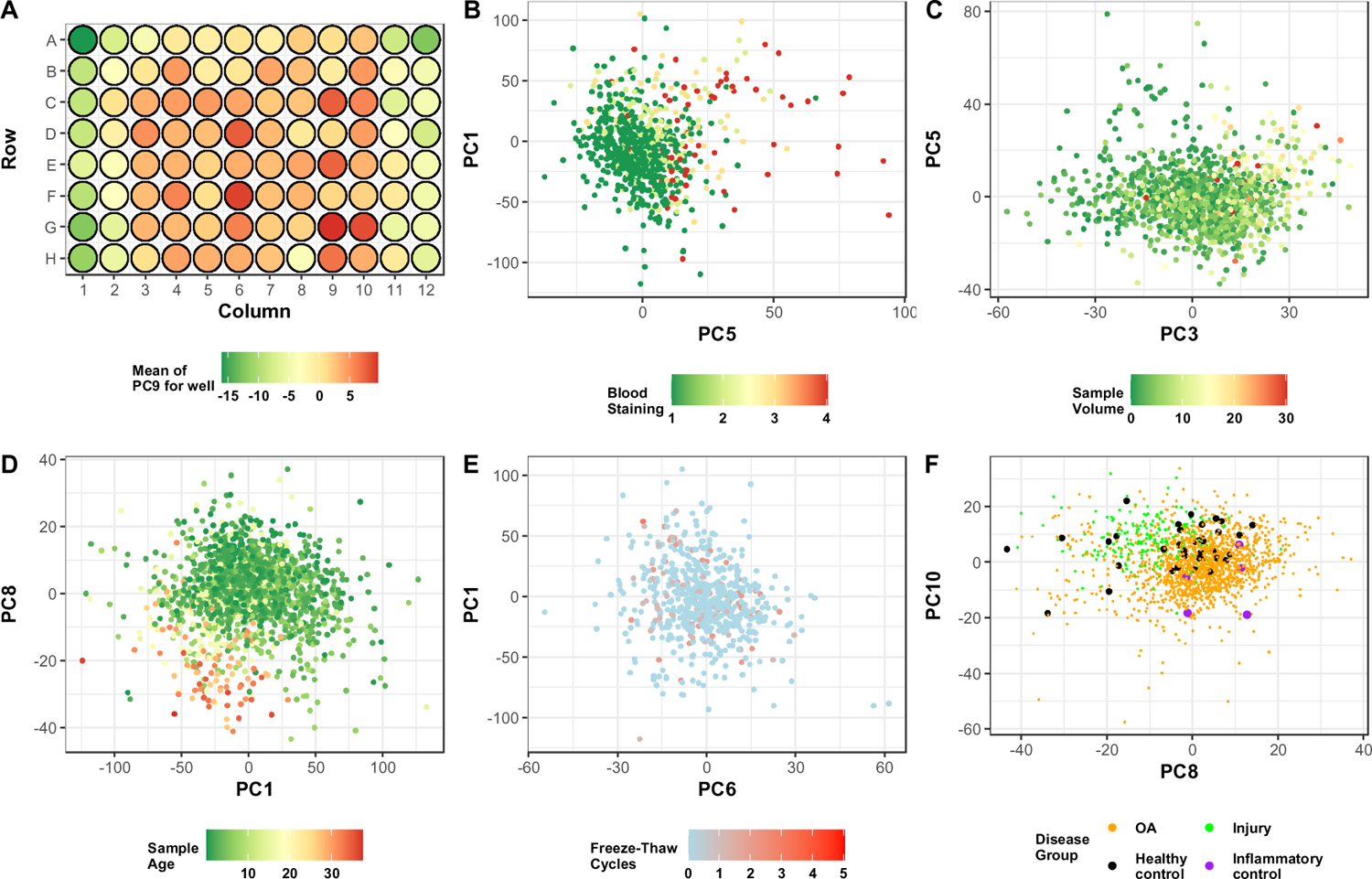
Visualisation of selected predefined confounders against select principal components of the batch corrected, filtered, IPS adjusted data. (A) The average value of PC9 (most strongly associated with plate position) by sample well position, (B-F) visualisation of the two PCs most strongly associated with each confounder, coloured by confounder value. Pre-defined confounders shown are (B) blood staining grade of sample after aspiration assessed by visual inspection, (C) volume of sample taken during aspiration, (D) age of the sample in years, measured from aspiration to sample processing at Oxford, (E) the number of times the sample was thawed and re-frozen before sample processing at Oxford, (F) the disease group of the sample (osteoarthritis [OA], acute knee injury [Injury], healthy control, inflammatory arthritis control).

Finally, we visualised the different diagnostic subgroups (OA, joint injury, inflammatory control, disease-free control) on UMAPs of the standardised, corrected and filtered data, with and without IPS adjustment (Figure 7A&B respectively). Both datasets showed clustering of knee injury and OA cases in overlapping but distinguishable areas of high-dimensional proteomic space, though the smaller groups (disease-free controls and inflammatory controls) were more evenly distributed. Inflammatory controls tended to segregate with acute knee injury samples. These patterns were also reflected at the PC level (Figure 6F, Figure S6).

**Figure 7.**
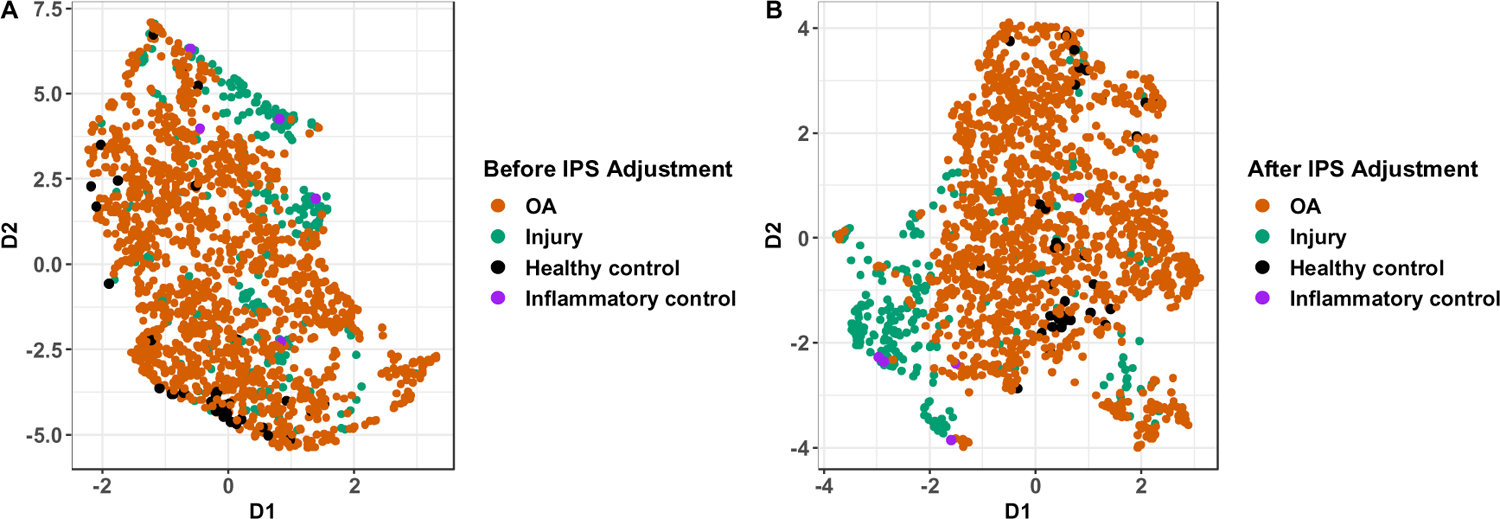
UMAP visualisation of two reduced dimensions (D1 and D2) of the top PCs of the log abundance data with (A) and without (B) IPS adjustment followed by filtering, coloured by disease group. These groups were osteoarthritis (OA, acute knee injury (injury), healthy controls, inflammatory arthritis controls.UMAP, Uniform Manifold Approximation and Projection.

## DISCUSSION

In this initial report from the STEpUP OA consortium, we describe a comprehensive evaluation of the overall performance of the SomaScan assay for knee SF for the first time. We address a series of data processing and analysis challenges that arise from proteomic quantification of SF using this technology. Based on our investigations, we propose an optimal standardisation procedure for SF and an assessment of the quality of the protein data using pre-defined approaches. Our aim was to justify the best approach to minimize technical variation while maintaining biological variation in the data, and thus to develop a pipeline that could be applied to downstream analyses within STEpUP OA and in subsequent proteomic analyses of SF by others.

We identified a number of technical confounders, which all affected, to a great or lesser extent, the data structure. These included factors that related to the SF sample e.g. its age, number of freeze-thaws, as well as potential confounders that could arise during the sample processing e.g. plate and date of processing. Because we had randomised the samples to and within plate, we were also able to identify plate position and laboratory processing batch as additional confounding factors. Each of these was controlled for by adjustment or filtering (either sample or SOMAmer). Whilst our intention is to perform the primary analysis of STEpUP OA only in spun SF sample data, we also included a number of unspun samples to test the generalizability of these to the larger dataset. In doing so we calculated that more stringent filtering would be needed when studying unspun, or mixed spun/unspun SF collections. We chose not to correct for blood staining (or HBA) or sample volume as we felt that these could reflect important biological variation.

The finding that a large proportion of variance in the data was driven by intracellular protein was unexpected. This could have arisen as a result of technical confounding following contamination of the SF samples by cells e.g. by a failure to remove cells fully by centrifugation, or by cell lysis of those cells at the time of aspiration e.g. by delay in spinning sample down. It could also be a true reflection of cellular turnover within the joint as part of the disease process e.g. of infiltrating immune cells or native connective tissues. It could also reflect protein carried within microvesicles that are known to be increased in joint disease and which drive biology within and between joint tissues [54, 55]. These possibilities are currently being explored. There is a concern that if the IPS reflects true biology, then adjusting for it may compress true signals within the data. This is consistent with the reduction in correlation with immunoassay seen in the injury samples. On the other hand, subtle structures within the data that reflect true molecular endotypes might be masked without removal of this signal. For this reason, the relevance of the IPS to clinical parameters and endotype clusters will be addressed alongside one another in the primary analysis of STEpUP OA.

Our data demonstrate that, when properly processed, SomaScan produces repeatable and accurate quantification of SF proteins which can (as a quality check, not a diagnostic one) broadly separate different clinical groupings. Its quantitation quality is at least comparable/superior to other ‘non-standard’ matrices e.g. urine/cerebrospinal fluid previously studied on this platform [56–58]. Our repeatability measures in pooled synovial fluid samples, with median %CVs of 11.25% for OA sample replicates and 12.42% for injury sample replicates, were higher than have been reported for plasma and serum samples measured using SomaScan technology, where %CVs of 5% or less are observed[59]. However, our correlation with immunoassays (median coefficient 0.81 for OA and 0.92 injury respectively) are as good or better than are typically observed in blood samples [60, 61]. Our data quality metrics are comparable with those based on mass spectrometry or other quantitative immunoassays, where %CVs of 10% or greater are recorded [62, 63]. Compared with these technologies our data has higher dynamic range and sensitivity (as the technology can assay proteins that are at very high as well as low abundance), noting only proteins on a pre-defined (though very large) protein list are included.

There are several limitations of this large study. The consortium collection was highly heterogeneous, gathered from seventeen different studies, varying in disease severity and phenotype, across several decades and from a number of countries without a unified pre-specified sample processing protocol. This made distinguishing technical and biological variation difficult (as was the case for the intracellular protein score, which could reflect either variation in joint biology or variation in sampling handling). A further limitation is that we analysed only a single matrix (SF) although this likely reflects activity in multiple joint tissues. The lack of paired cartilage/synovium/bone in STEpUP OA prevents us performing a direct integrated analysis using RNAseq, for example, although it may be possible to extrapolate this from other existing datasets.

Other proteome-wide technologies (such as LC-MS/MS, OLINK[64]) could provide further validation on protein patterns within OA SF. Paired plasma is also available for many individuals in STEpUP OA but is yet to be analysed. From previous experience we would predict that this matrix would show low concordance with SF [41, 65]. Others have used SomaScan to explore the plasma proteome in OA, identifying diagnostic and prognostic biomarkers, though this study did not study paired SF samples[34].

In summary, we present an evidence-based methodology pipeline for large scale proteomic analysis on the SomaScan platform of SF, which has the potential to be a critical matrix for discovery science and clinical translation in OA. Our next step, the primary analysis of this dataset, seeks to answer definitively whether there are distinct discernible molecular endotypes in this common, yet poorly understood disease.

## Author contributions

Conception and Design: TLV, FEW, LJD, PH, RAM, DP, SL, SB, LSL, AS, CTA, DF, BDMT, MK, TJW, DAW, AMV. Analysis and interpretation of data: YD, TAP, LJD, FEW,TLV, PH, RAM, JM, SB, BDMT, LB. Drafting Article: TLV, TAP, YD, LJD, FEW. Critical revision of article: all authors. Final Approval: all authors.

## Data Availability

In accordance with the STEpUP OA Consortium Agreement and the Data Access and Publication Group, protein and clinical data will be available for bone fide research relating to osteoarthritis through an application to the STEpUP OA Data Access and Publication group once the discovery and replication analyses are in press and if it does not infringe patent position. This may be subject to an access fee (to be confirmed).

## Acknowledgements

We would like to express our gratitude and thanks to all cohorts and their participants who contributed samples to STEpUP OA. We are grateful for the support from Floris Lafeber and Simon Mastbergen (Utrecht Medical Centre). This work was also supported by the NIHR Oxford Biomedical Research Centre (BRC) and the NIHR Nottingham BRC. The views expressed are those of the authors and not necessarily those of the NHS, the NIHR or the Department of Health. Tissue samples and/or data obtained from the Oxford Musculoskeletal Biobank were collected with informed donor consent in full compliance with national and institutional ethical requirements, the UK Human Tissue Act, and the Declaration of Helsinki (HTA Licence 12217 and Oxford REC C 09/H0606/11). We thank the Oxford Knee Surgery Team including Andrew Price, William Jackson and Nicholas Bottomley and our centre tissue coordinators Louise Hill and Katherine Groves who coordinated this study. We thank Charlotte Kerr for her administrative support of the consortium at large.

<u>The STEpUP OA Consortium author block</u> includes: University of Nottingham: Ana M. Valdes, David A. Walsh, Michael Doherty, Vasileios Georgopoulos; Lund University: Staffan Larsson, L. Stefan Lohmander, André Struglics; University of Cambridge: Brian D.M. Tom, Laura Bondi; University of Toronto: Mohit Kapoor, Rajiv Gandhi, Anthony Perruccio, Y. Raja Rampersaud, Kim Perry; University of Manchester: Tim Hardingham, David Felson; University of Oxford: Tonia L. Vincent, Thomas A. Perry, Luke Jostins-Dean, Yun Deng, Vicky Batchelor, Jennifer Mackay-Alderson, Gretchen Brewer, Rose M. Maciewicz, Brian Marsden, Nigel K. Arden, Philippa Hulley, Andrew Price, Stefan Kluzek, Megan Goff, Vinod Kumar, James Tey; Imperial College London: Fiona E. Watt, Andrew Williams, Artemis Papadaki; University College Maastricht: Tim J. Welting, Pieter Emans, Tim Boymans, Liesbeth Jutten, Marjolein Caron, Guus van den Akker; University of Western Ontario: C. Thomas Appleton, Trevor B. Birmingham, J. Daniel Klapak; Biosplice: Sarah Kennedy, Jeymi Tambiah; Fidia: Devis Galesso, Nicola NK; SomaLogic: Joe Gogain, Darryl Perry, Anna Mitchel, Ela Zepko; Novartis: Sophie Brachat, Joanna Mitchelmore, Juerg Gasser, Lori Jennings; UCB: Waqar Ali.

TLV directs the Centre for OA pathogenesis (grant numbers 21612 and 20205) and has additional grant support from Versus Arthritis, the European Research Council, the Medical Research Council and FOREUM. LJD is supported by a Wellcome trust fellowship grant 208750/Z/17/Z and Kennedy Trust for Rheumatology Research for the present manuscript. LJD is also supported by grants from the MRC and the Helmsley Charitable Trust. FEW is supported by a UKRI Future Leaders Fellowship (MRC number: MR/S016538/1 and MR/S016538/2). FW, NKA and SK are members of the Centre for Sport, Exercise and Osteoarthritis Research Versus Arthritis (grant number 21595). MK is supported by grants from CIHR, NSERC, The Arthritis Society Canada, Krembil Foundation, CFI, Canada Research Chairs program, and has received support from the University Health Network Foundation, Toronto for the present manuscript. TJW is supported by grants from NWO-TTW Perspectief (#P15-23), Stichting de Weijerhorst and ReumaNederland (LLP14) for the present manuscript, and is a shareholder of Chondropeptix BV. BDMT is supported through the United Kingdom Medical Research Council programme (grant MC UU 00002/2). For the purpose of open access, the authors have applied a Creative Commons Attribution (CC BY) license to any Author Accepted Manuscript version arising. LB is supported by grants from Kennedy Trust for Rheumatology Research (grant number 171806) and UK Medical Research Council (grant MC UU 00002/2). DAW is supported by grants from Pfizer Ltd, UCB Pharma, Orion Corporation, GlaxoSmithKline Research and Development, and Eli Lilly and Company, Versus Arthritis, UKRI, Nuffield Foundation.

## Ethical Approval

The ethical approval reference numbers for individual participating cohorts are provided in Table S1. In addition, a University CUREC approval was granted for the study (details in Methods).

## Supplementary Data

Code availability https://github.com/dengyun-git/STEpUp_QC_Paper. For access to primary data used in this analysis, see Data Access section.

## Patient and Public Involvement Statement

People with lived experience of osteoarthritis have been involved in the design of this project. A patient research panel was involved in discussing and inputting on the STEpUP OA project in February 2020 (invited to the Centre for Osteoarthritis Pathogenesis Versus Arthritis in Oxford, as part of its involvement activities). Aspects relevant to the development of the project were further discussed with the panel in July 2022. The working groups for the consortium include one focused on patient involvement and engagement. A lay summary is included in the appendix of our publicly available analysis plan. A short video about the project was produced and is available on our website: https://www.kennedy.ox.ac.uk/oacentre/stepup-oa. In addition, the various constituent cohorts contributing to STEpUP OA also typically have lay or patient members on their steering committees.

---

**Figure S1.** Consortium structure, as working groups. Distinct working groups oversaw key activities according to pre-defined Terms of Reference (available on request). TV, Tonia Vincent; FW, Fiona Watt; AV, Ana Valdes; LJD, Luke Jostins-Dean; RM, Rose Maciewicz.

**Figure S2.** Assessment of assay repeatability using pooled samples of synovial fluid from participants with (A) knee OA and (B) acute knee injury, measured by the coefficient of variation (%CV). These include the repeatability of the standard processed pooled samples included on every plate (‘Sample Repeats’), pooled samples which had been repeatedly freeze-thawed (‘Freeze Thaw’) prior to processing and an OA pool aliquot that had been freshly enzyme digested with stored hyaluronidase during each of the 2nd, 3rd and 4th tranches of sample processing (done for the OA pool only) (‘Reprocessed’). Dotted vertical lines show the maximum %CV for 80% of proteins for each group.

**Figure S3.** Top 2 principal components of (A) non-IPS adjusted and (B) IPS adjusted log RFU of the 18 pairs of centrifuged (spun) and non-centrifuged (unspun) SF samples. Samples are coloured by spin status and paired samples are linked by lines. Measures of differential abundance (Cohen’s d) and Pearson correlation coefficient (rho) between spun and unspun samples for (C) non-IPS adjusted and (D) IPS adjusted log RFU. Samples are coloured depending on their significance (Benjamini-Hochberg adjusted p< 0.05) on the two measures: Different Means corresponds to a significant difference in means in a paired t-test and Correlated corresponds to a significant correlation in a Pearson correlation test. IPS, intracellular protein score; PC, principal component; SF, synovial fluid; RFU, relative fluorescence unit.

**Figure S4.** Boxplots showing the correlation between visual blood staining grade of SF at the time of sample collection and the blood analyte, HBA, in non-IPS adjusted data in (A) all samples, (C) OA samples and (E) acute knee injury samples, and in IPS adjusted data in (B) all samples, (D) OA samples and (F) acute knee injury samples. Spearman correlation coefficients measuring rank-based correlation considering visual blood staining as an ordinal variable are shown. 443 OA samples had blood staining grade 1 (no blood detected, 75% among the 588 total samples with blood staining records). HBA, haemoglobin A; IPS, intracellular protein score; SF, synovial fluid.

**Figure S5.** (A) Assessment of assay repeatability after optimised quality control procedures measured using the cumulative distribution of the coefficient of variation (%CV) on pooled OA samples (OA Sample Repeats) and pooled acute knee Injury samples (Injury Sample Repeats) separately. 80% of proteins had a %CV less than 16.85% and 17.57% in the OA and acute knee injury pools (blue and red dotted lines respectively). (B) The proportion of variation that was estimated to be non-technical, measured by R ^2^ for OA and acute knee injury sample repeats separately. 80% of proteins had R ^2^ values greater than 88.27% and 84.33% in the OA and knee injury pools (blue and red dotted lines respectively).

**Figure S6.** Visualisation of pre-defined technical confounders by select principal components of the (A) non-filtered IPS adjusted (B) filtered non-IPS adjusted and (C) non-filtered non-IPS adjusted data. Visualisation of the two PCs most strongly associated with each confounder (colours correspond to confounder value). Confounders include plate position (mean of PC8), blood staining grade of sample (which was performed immediately after aspiration from the joint by visual inspection), volume of sample taken during aspiration, age of the sample in years from aspiration to processing, the number of times the sample had been thawed and re-frozen, the disease group of the sample (knee OA, acute knee injury, healthy control, inflammatory arthritis control). The association between each PC and confounder is shown in the Table S7.

**Figure S7.** Pairwise scatter plots (off-diagonal) and histograms (diagonal) of the top five principal components of standardised log abundance, (A) before and (B) after batch correction for plate and bimodal signal status, coloured by bimodal signal status. Batch correction effectively removed the effect of bimodal signal status on the top PCs.

