## Supplementary Tables for "Methodological development of molecular endotype discovery from synovial fluid of individuals with knee osteoarthritis: the STEpUP OA Consortium"

| Cohort Name and ethical approval number | City, Country | QC Sample Grouping^1^ | Disease Type/Severity/Surgical procedure | Total Samples Contributed^2^  N (n) | Samples submitted for QC^3^N (n) | Sex^4^, female n (%) | Age^4^, median (IQR) | Ordinal KL grade^4,5^, median (IQR) |
| --- | --- | --- | --- | --- | --- | --- | --- | --- |
| *KANON*  (LU 535-01, ISRCTN84752559) | Lund, Sweden | Acute knee Injury | Anterior cruciate ligament tear | 72 (44) | 72 (44) | 12 (16.7) | 24 (7) | 0 (0) |
| *KICK*  (10/H0805/39) | Oxford, UK | Acute knee Injury | Acute knee injury | 140 (140) | 140 (140) | 26 (18.6) | 25 (9.5) | 1 (2) |
| *OxKIC*  (14/WA/1108) | Oxford, UK | Acute knee Injury | Anterior cruciate ligament and/or meniscal injury | 33 (33) | 33 (33) | 8 (24.2) | 32 (13) | 0 (0) |
| *COASt*  (10/H0604/91) | Oxford, UK | OA | Knee arthroplasty | 20 (20) | 20 (20) | 13 (65) | 73 (13) | - |
| *MJC*  (2017-0183) | Maastricht, Netherlands | OA | Knee arthroplasty | 237 (219) | OA: 218 (200) | 115 (52.8) | 69 (14) | 3 (1) |
| *Distraction study*  (160/D; NL51539.041.15) | Utrecht, Netherlands | OA | Joint distraction | 38 (20) | 38 (20) | 13 (34.2) | 55 (8) | 3 (1) |
| *EARLY OA*  (REB 16-5969 AE) | Toronto, Canada | OA | Non-advanced disease | 43 (43) | 43 (43) | 19 (44.2) | 58 (13) | 2 (0) |
| *LEAP OA*  (16-5759 BE) | Toronto, Canada | OA | Knee arthroplasty | 339 (339) | 338 (338) | 189 (55.9) | 66 (12) | - |
| *LUMEN*  (LU 49-98) | Lund, Sweden | OA | Varying severity | 32 (32) | 32 (32) | 9 (28.1) | 62 (16) | - |
| *LUMEX*  (LU 361-00) | Lund, Sweden | OA | Varying severity | 67 (67) | 67 (67) | 34 (50.8) | 64 (10) | 3 (1) |
| *DBOARD*  (14/EM/0013) | Nottingham, UK | OA | Varying severity | 46 (46) | 46 (46) | 26 (56.5) | 68.5 (9) | 2 (3) |
| *MenTOR*  (15/SC/0551) | Oxford, UK | OA | Varying severity | 75 (75) | 75 (75) | 21 (28) | 47 (11) | 2 (1) |
| *NBS GOAL*  (EC2/06) | Nottingham, UK | OA | Varying severity | 112 (112) | OA: 74 (74) | 33 (44.6) | 67 (12) | 3 (1) |
|  |  | Non-OA controls | No radiographic disease |  | Non-OA controls: 31 (31) | 8 (25.8) | 65 (17) | 0 (0) |
| *OMB*  (09/H0606/11)  *MMOA*  (07/H0706/81) | Oxford, UK | OA / non-OA controls / injury | Arthroplasty | 199 (199) | OA: 194 (194) | 105 (54.1) | 69 (12) | - |
|  |  |  |  |  | Non-OA control(s): 1 (1) | 1 (100) | 46 (-) | - |
|  |  |  |  |  | Injury: 1 (1) | 0 | 29 (-) | - |
| *Molecular Pathways*  (20/EM/0065) | Nottingham, UK | OA | Varying severity | 45 (45) | 45 (45) | 27 (60) | 67 (10) | 2 (1) |
| *WebEx*  (18/EM/0154) | Nottingham, UK | OA | Varying severity | 20 (20) | 20 (20) | 13 (65) | 66 (13.5) | 2 (1) |
| *WOREO*  (REB 109255) | London, Canada | OA | Varying severity | 245 (207) | 236 (204) | 135 (57.2) | 65 (15) | 3 (2) |
| *Other:*  *NBS GOAL*  (EC2/06)  *Understanding pathogenesis of OA* (16/LO/1351)  Molecular Mechanisms  (07/H0706/81) | Nottingham, UK  Oxford, UK  Oxford, UK | OA / non-OA controls / inflammatory controls | - | 17 (15) | OA: 5 (5) | 2 (40) | 81 (3) | - |
|  |  |  |  |  | Non-OA controls: 5 (5) | 2 (40) | 64.5 (21.5) | - |
|  |  |  |  |  | Inflammatory controls: 5 (5) | 3 (60) | 47 (9) | - |

**Table S1.** ***Characteristics of Cohorts and Participant Samples.***

^1^ QC sample grouping: a broad description of the participant’s membership, attributed by cohort-level information at baseline, of one of the following groups:

Knee OA, acute knee joint injury, non-OA (healthy) controls, inflammatory arthritis controls.

^2^N shown is the total number of SF samples contributed to STEpUP OA analysis by each cohort (total = 1780 samples) before sample exclusions prior to inclusion into the quality control (QC) analysis.

^3^In parentheses, n shown is the number of samples passing QC for each cohort (total = 1746 samples). 34 of 1780 (1.9%) samples were excluded from the QC analysis: 10 samples were excluded at processing stage, 18 samples were of insufficient volume to be processed by SomaLogic and 6 were contralateral knee samples from same participant to be used in later analyses.

^4^Demographic and basic clinical data for those samples passing QC analysis (1746 samples) at the time of SF sample collection are summarized.

^5^For the LUMEN cohort, whilst radiographic scores were available, these were not ordinal KL grades so not reported here.

Abbreviations: OA (osteoarthritis), KL (Kellgren and Lawrence), UK (United Kingdom), QC (Quality Control), Nonsurgical versus Surgical Treatment Study (KANON), Knee Injury Cohort at the Kennedy (KICK), Oxford Knee Injury Cohort (OxKIC), Clinical Outcomes in Arthroplasty Study (COASt), Lund Meniscus cohort (LUMEN), Osteoarthritis Biomarker Follow up study (D-BOARD consortium), MJC, Maastricht Joint Collection, Meniscal Tear and Osteoarthritis Risk (MenTOR), Nottingham Biomarker Study Genetics of Osteoarthritis and Lifestyle (NBS GOAL), Oxford Musculoskeletal Biobank (‘OA pathogenesis’ project) (OMB), Molecular Mechanisms in OA (MMOA), Western Ontario Registry for Early Osteoarthritis (WOREO) Knee Study.

| **Applied to** | **Field** | **Description** | **Coding** |
| --- | --- | --- | --- |
| **Quality Control  and Downstream Analysis** | **sf_iknee_sample_id_number** | The STEpUP OA Sample Identification Number (SIN) | string |
|  | **stepup_id** | The STEpUP OA Participant Identification Number(PIN) | string |
| **Quality Control** | **age_sampling** | Patient age at the time sample was taken (to the nearest year) | integer (NA=missing) |
|  | **sl_plate_id** | Identification (ID) of plate the sample was run on | string |
|  | **sl_plate_run_date** | Date that the sample was run | string (“dd-mm-yyyy”) |
|  | **sl_plate_position** | Position of the sample on the 96-well plate | string (“XN”, where X is row letter and N is the column number) |
|  | **sl_scanner_id** | ID of the SomaScan scanner that the sample was read on | string |
|  | **sl_tranche_number** | Shipment tranche in which sample was run | {1 = tranche 1, 2 = tranche 2} |
|  | **sl_bimodal_signal** | The technical bimodal signal, strongly correlated with processing batch, used to batch-correct the data. | {bimodal1, bimodal2 - arbitrary labels for the two groups. NA = missing} |
|  | **sf_iknee_proc_batch** | Batch number for index knee sample | Integer (NA = missing) |
|  | **sf_iknee_proc_order** | Processing order number for knee samples | Integer (NA = missing) |
|  | **sf_iknee_proc_treat_date** | Date sample was hyaluronidase-treated by Oxford | Text (dd-mm-yyyy) |
|  | **sf_iknee_qc_group** | Patient grouping (OA, acute knee injury or control) at baseline | {0 = OA, 1 = Joint injury, 2 = healthy control, 3 = inflammatory control, NA = missing} |
|  | **cohort_name** | Cohort ID (an arbitrarily chosen integer assigned to each cohort) | integer |
|  | **sex** | Patient sex at baseline (as defined by individual cohort collectors) | {m = male, f = female, NA = missing} |
|  | **sample_age** | Time between date of sample collection and date of STEpUP OA sample processing for the index knee (years) | float (years) (NA = missing) |
|  | **sf_iknee_volume** | Total SF volume collected (ml) | float (ml) |
|  | **sf_iknee_prev_freeze_thaw** | Whether the sample had been freeze-thawed prior to STEpUP OA sample processing | {0 = No, 1 = Yes, NA = Unknown} |
|  | **sf_iknee_freezethaw_cycles** | Number of freeze-thaw cycles (if known) | integer (NA=missing) |
|  | **sf_iknee_freezethaw_spec** | Whether the sample  has been freeze-thawed less than, or greater to or equal to five times | {0 = <5, 1 = ≥5, NA = missing} |
|  | **sf_iknee_bloodstaining** | Grading of SF bloodstaining prior to centrifugation (if known). Scale of 1-4, with larger numbers corresponding to greater degrees of blood staining (by visual inspection) | {1 = None, 2 = Mild, 3 = Moderate, 4 = Severe, NA = Not known} |
|  | **sf_spun_vs_unspun** | Indicator for whether the sample was centrifuged prior to being received at Oxford | 0 = unspun, 1 = spun, 2 = not known |
| **Downstream Analysis (Discovery Analysis & Replication Analysis)** | **Cohort name** | Cohort ID (an arbitrarily chosen integer assigned to each cohort) | integer |
|  | **Disease allocation** | Patient grouping (OA, acute knee injury or control) at baseline. Note that this estimate of disease was based primarily on the inclusion criteria of the individual cohorts, not at individual level | {0 = OA, 1 = acute knee injury, 2 = healthy control, 3 = inflammatory control, NA = missing} |
|  | **Age** | Patient age at the time sample was taken (to the nearest year) | integer (NA = missing) |
|  | **Sex** | Patient sex at baseline (as defined by individual cohort collectors) | {m = male, f = female, NA = missing} |
|  | **BMI** | Patient body mass index at the time the sample was taken (calculated from provided height and weight or directly provided by cohort collector, in that order of preference) | float (kg/m^2) |
|  | **Ordinal KL grade (worst affected compartment)** | Kellgren-Lawrence (KL) grade of radiographic severity at time of sampling | {0 = grade 0 (none), 1 = grade 1 (doubtful), 2 = grade 2 (minimal), 3 = grade 3 (moderate), 4 = grade 4 (severe), NA = Missing OR Not Known} |
|  | **Binary indicator for the presence/absence of radiographic knee OA** | Flag indicating whether the sample was taken from a patient with radiographic OA in the index knee, defined as a KL grade greater or equal to two at time of sampling | {0 = No (i.e. KL < 2), 1 = Yes (i.e. KL ≥2, NA = Missing OR Not Known} |
|  | **Binary indicator for the presence of advanced stage radiographic knee OA (KL scores 3-4)** | Flag indicating whether the sample was taken from a patient with advanced radiographic OA in the index knee, defined as a KL grade greater or equal to three at time of sampling | {0 = No (i.e. KL < 3), 1 = Yes (i.e. KL ≥3), NA = Missing OR Not Known} |
|  | **Smoking history** | Flag indicating whether the patient was a current or past smoker at the time of the baseline sample | {0 = No (i.e. never smoked), 1 = Yes (i.e. current smoker or past smoker), NA = missing or not available} |
|  | **baseline** | Flag indicating whether this sample is a baseline sample (each individual has one baseline sample) | {0 = No, 1 = Yes} |
|  | **Harmonised Knee Pain Score** | Binary flag indicating whether experienced knee pain is outside of the Patient Acceptable Symptom State (PASS) at the time of sampling (calculated from the KOOS pain subscale, the WOMAC pain subscale or knee VAS (knee-specific NRS/VAS or painDETECT VAS, in order of preference). | {0 = No (acceptable pain), 1 = Yes (unacceptable pain), NA = missing or Not Available} |
|  | **Harmonised Patient Reported Outcome Measure (PROM)** | The specific patient reported outcome measure used to derive a harmonised knee pain score | {1 = KOOS, 2 = WOMAC, 3 = Knee specific VAS/NRS,  4 = PainDETECT VAS, NA=missing} |
|  | **KOOS pain score** | KOOS pain subscore (calculated from full KOOS questionnaire results, or from combined subscore provided by cohort collectors, in that order of preference). Scale of 0-100, where 0 is the worst possible pain recordable. | Float |
|  | **WOMAC pain score** | WOMAC pain subscore (calculated from full WOMAC questionnaire results, or from combined subscore provided by cohort collectors, or derived from full KOOS questionnaire results, in that order of preference). Scale of 0-100, where 100 is the worst possible pain recordable. | Integer |
|  | **Knee-specific numeric rating score (NRS)** | Patient reported knee pain on a Numeric Rating Scale (0-10), where 10 is the worst pain imaginable | Float |
|  | **PainDETECT numeric rating score (NRS)** | Patient reported average pain score (over the last 4 weeks) from the painDETECT questionnaire. Scale of 0-10, where 10 is the worst pain imaginable | Integer |

**Table S2.** ***Core Clinical Phenotype Data used for Quality Control and Downstream Analyses.***

These data include sample information used to carry out quality assessment, as well as clinical phenotype data required for the downstream Discovery and Replication analyses. ‘Plate’ refers to that used to assay the sample at SomaLogic. ‘Batch’ refers to the membership of a group of sessional processing by the Oxford Lab. ‘Tranche’ relates to overall larger groupings of sample processing (four in total) carried out by the Oxford Lab, where processed samples from each group were shipped together to SomaLogic. Note that details of various pre-defined knee pain measures and harmonised scores are given here, but their analysis is not reported in this manuscript (variables were predefined for the discovery and replication analysis). This is also true for some of the measures of radiographic severity (KL grade is included here). Abbreviations: WOMAC, Western Ontario and McMaster Universities Osteoarthritis Index; KOOS, The Knee injury and Osteoarthritis Outcome Score; NRS, Numeric rating score; VAS; visual analogue scale; PROM, patient reported outcome measure; SF, synovial fluid; BMI, Body Mass Index.

| **Target** | **Target Full Name** | **Type of Assay** |
| --- | --- | --- |
| **TIMP-1** | Tissue Inhibitor of Metalloproteinase-1 | MSD Ultra-Sensitive |
| **MMP-3** | Matrix Metalloproteinase-3 (Stromelysin-1) | MSD Ultra-Sensitive |
| **IL-6** | Interleukin-6 | MSD V-PLEX |
| **IL-8** | Interleukin-8 | MSD V-PLEX |
| **MCP-1** | Monocyte Chemoattractant Protein-1 (C-C motif chemokine 2) | MSD V-PLEX |
| **FGF2** | Fibroblast growth factor 2 | MSD V-PLEX |
| **Activin A** | Activin A | R&D Quantikine ELISA |
| **TGFβ1** | Transforming growth factor Beta-1 | R&D Quantikine ELISA (active and latent combined) |
| **TSG-6** | Tumor necrosis factor-inducible gene 6 | MSD inhouse assay (U plex) |

**Table S3. *Summary of Proteins Measured by Immunoassay used to assess accuracy of SomaScan Data.***

Nine protein targets included on the SomaScan platform (V4.1) had been previously measured using immunoassay of a subset of the synovial fluid samples (albeit not hyaluronidase treated), by either conventional sandwich ELISA (R&D) or by electrochemiluminescent assay (MSD) ("Type of Assay") following manufacturer’s instruction, and these data were used to evaluate the degree of agreement with the SomaScan data. Manufacturers were: MSD, Mesoscale Discovery, Rockville and R&D, R&D Systems, Minneapolis, both US.

| **Filter label** | **Filter Description** | **Description** | **Applies To** | **Excluded**  **Proteins/Samples**  **within**  **Non-IPS adjusted data** | **Excluded Proteins/Samples within**  **IPS adjusted data** |
| --- | --- | --- | --- | --- | --- |
| **NONHUMAN** | Non-human proteins | Non-human or control proteins | Proteins | 307 | 307 |
| **OA_REPO** | Reproducibility in OA pool | R^2^ < 0.5  (non-technical variation less than 50%) | Proteins | 485 | 485 |
| **INJ_REPO** | Reproducibility in acute knee injury pool | R^2^ < 0.5  (non-technical variation less than 50%) | Proteins | 252 | 252 |
| **FREEZETHAW_CONFOUND** | Associated with number of freeze-thaw cycles | ANOVA p < 0.05/7289  (conditional on cohort) | Proteins | 56 | 212 |
| **SAMPLEAGE_CONFOUND^1^** | Associated with sample age | ANOVA p < 0.05/7289  (conditional on cohort) | Proteins | 77 | 229 |
| **BIMODAL_CONFOUND** | Associated with bimodal signal | ANOVA p < 0.05/7289 | Proteins | 96 | 72 |
| **SOMASCAN_FAIL** | SomaLogic inhouse QC | Hybridization Scale Factor > 2.5 | Samples | 2 | 2 |
| **LOD_SAMPLE^2^** | Limit of detection | 25% of proteins below/above  limit of detection | Samples | 12 | 12 |
| **TOTPROT_OUTLIER** | Total protein outliers | >5 SDs from mean | Samples | 9 | 9 |
| **PCA_OUTLIER** | PCA outliers | >5 SD from combined centre on top PCs | Samples | 15 | 15 |
| **Total (number after filtering /number before filtering)** | | | Proteins | 6558/7596  (86.33%) | 6290/7596  (82.81%) |
|  |  |  | Samples | 1720/1746  (98.51%) | 1720/1746  (98.51%) |

**Table S4.** ***Summary of Sample and Protein Filters.***

Details of the filters applied to the batch corrected, non-IPS adjusted data or IPS adjusted data, including thresholds used and number of samples or proteins (SOMAmers) removed by each filter. The final row gives the total number of proteins and samples remaining in the two datasets after filtering, to be used in downstream analyses. Note that proteins and/or samples removed were not mutually exclusive across filters. Abbreviations: IPS, intracellular protein score; PCA, Principal Component Analysis; SD, standard deviation. P values were Bonferroni corrected.

^1^Sample age was defined as the time from SF collection to time of SF processing at Oxford. ^2^Lower limit of detection (lLoD) was defined as the median concentration of the buffers plus 4.9 times the median absolute deviation of the buffers, and upper limit of detection (uLoD) was defined as 80,000 RFU, both as recommended by SomaScan platform.

| **IPS Adjustment** | **Predictor** | **Regression Coefficient** | **P-value** |
| --- | --- | --- | --- |
| **Non-IPS Adjusted Data** | ***Average protein abundance (log mean abundance)*** | ***-0.056*** | ***<2.23e-308*** |
|  | ***Non-secreted Nuclear protein (Y/N)^1^*** | ***0.033*** | ***1.64e-09*** |
|  | ***Non-secreted protein (Y/N) ^1^*** | ***0.051*** | ***3.98e-10*** |
|  | Monocyte protein (Y/N)^2^ | 0.033 | 0.077 |
|  | Neutrophil protein (Y/N)^2^ | 0.015 | 0.48 |
|  | Macrophage protein (Y/N)^2^ | -0.00044 | 0.98 |
| **IPS Adjusted Data** | ***Average protein abundance (log mean abundance)*** | ***-0.0080*** | ***5.8e-16*** |
|  | Non-secreted Nuclear protein (Y/N)^1^ | 0.012 | 0.25 |
|  | Non-secreted protein (Y/N) ^1^ | -0.019 | 0.23 |
|  | Monocyte protein (Y/N)^2^ | 0.038 | 0.30 |
|  | Neutrophil protein (Y/N)^2^ | -0.059 | 0.17 |
|  | Macrophage protein (Y/N)^2^ | 0.021 | 0.53 |

**Table S5. *Predictors of the strength of correlation between protein abundance and PC1.***

Multiple linear regression results (coefficient and p-value) for the effect of a variety of protein-level factors on the correlation between each protein signal intensity and PC1. Protein abundance is calculated as the standardized RFU for each protein adjusted by the protein's dilution factor used in the SomaScan assay (the "dilution bin"). The dependent variable was the Pearson correlation between PC1 and protein signal intensity on a log scale. The independent variables were the average protein abundance on a log scale in SF, and flag indicating whether a protein was classified as a non-secreted protein (i.e. a protein not predicted to be secreted by the protein atlas) or a marker protein for monocytes, macrophages or neutrophils (taken from <https://panglaodb.se/index.html>), three common infiltrating immune cell types in the synovial joint. Results of log abundances and PC1 from the non-IPS adjusted and the IPS adjusted data are shown separately. Rows in bold italic show significant (p< 0.05) predictors, though all findings were also significant after Bonferroni correction for multiple testing. Abbreviations: IPS, intracellular protein score; SF, synovial fluid.

| **Protein Target** | **Activin A** | **FGF2** | **IL6** | **IL8** | **MCP1** | **MMP3** | **TGFb1** | **TIMP1** | **TSG6** |
| --- | --- | --- | --- | --- | --- | --- | --- | --- | --- |
| **Protein Full Name** | Activin A | Fibroblast growth factor 2 | Interleukin-6 | Interleukin-8 | Monocyte Chemoattractant Protein-1  (C-C motif chemokine 2) | Matrix Metalloproteinase- 3 (Stromelysin-1) | Transforming growth factor Beta-1 | Tissue Inhibitor of Metalloproteinase-1 | Tumor necrosis factor-inducible gene 6 |
| **OA** | 0.160  (0.324) | 0.250  (0.119) | 0.085  (0.604) | 0.381  (0.015) | 0.160  (0.323) | 0.112  (0.491) | 0.008  (0.964) | 0.204  (0.207) | 0.261  (0.104) |
| **Injury** | 0.481  (3.73e-2) | 0.683  (1.26e-3) | 0.585  (8.49e-3) | 0.639  (3.23e-3) | 0.598  (6.81e-3) | -0.073  (0.765) | 0.806  (3.06e-5) | 0.377  (0.112) | NA |

**Table S6. *Correlation between Intracellular Protein Score and protein abundance measured by Immunoassay.***

Nine proteins measured on SomaScan were also measured on conventional immunoassay (see Supplementary Table S3 for details of assays used). This table shows the correlation (correlation coefficients, with p-values in parentheses, both derived from Pearson correlation testing) between intracellular protein score and concentrations of key proteins measured on immunoassay, stratified by osteoarthritis (OA) and acute knee injury (Injury) samples. [Activin A, FGF2, IL6, IL8, MCP1, TGFb1 concentrations showed significant correlation with intracellular protein score. The estimated correlation coefficients were higher in injury samples compared with OA samples for 7 out of 8 proteins with data in both groups]**.**

| **Proteomic Data** | **Confounders** | **Plate ID** | **Disease Group** | **Plate Position** | **Plate Run Date** | **Sample Processing Tranche** | **Sample Previous Freeze Thaw**  **(Y/N)** | **Sample Freeze Thaw Cycles** | **Sample Processing Batch** | **Date of Hyaluronidase Treatment** | **Sample Age** | **Sample Volume** | **Visual Blood Staining** |
| --- | --- | --- | --- | --- | --- | --- | --- | --- | --- | --- | --- | --- | --- |
|  | **PC (variation explained)** |  |  |  |  |  |  |  |  |  |  |  |  |
| **Standardised +**  **Bimodal Signal Adjustment +  IPS adjustment + Filtering** | PC1 (~17.0%) | 1.00E+00 | **8.98E-16** | **6.81E-05** | 9.48E-01 | **7.74E-04** | 1.96E-01 | 1.18E-01 | **1.81E-03** | **1.30E-04** | **2.81E-04** | **9.76E-04** | **5.97E-13** |
|  | PC2 (~6.9%) | 1.00E+00 | **1.01E-19** | 1.02E-02 | 9.44E-01 | 1.73E-02 | 2.32E-02 | 1.20E-01 | **1.61E-09** | **1.17E-04** | 3.55E-03 | **3.26E-04** | 7.44E-01 |
|  | PC3 (~4.8%) | 1.00E+00 | **2.58E-23** | **5.20E-04** | 9.74E-01 | **4.41E-11** | **2.74E-05** | 3.27E-01 | **4.32E-39** | **3.19E-15** | **1.28E-03** | **9.95E-23** | 5.39E-03 |
|  | PC4 (~4.0%) | 1.00E+00 | 3.24E-02 | **1.02E-05** | 9.62E-01 | 2.70E-01 | 8.02E-01 | 5.80E-01 | **4.33E-16** | **3.17E-07** | **2.67E-03** | 5.66E-01 | 1.30E-01 |
|  | PC5 (~3.4%) | 1.00E+00 | **8.18E-28** | 9.56E-02 | 9.72E-01 | 2.51E-01 | 6.68E-01 | 2.90E-01 | **3.06E-15** | **2.07E-04** | 9.63E-01 | **2.77E-06** | **1.70E-34** |
|  | PC6 (~2.9%) | 1.00E+00 | **1.20E-05** | **3.35E-03** | 9.82E-01 | **3.33E-06** | **9.88E-09** | **2.58E-08** | **1.25E-32** | **3.85E-21** | 9.08E-03 | **7.15E-05** | 5.10E-03 |
|  | PC7 (~2.4%) | 1.00E+00 | **3.72E-07** | **4.21E-37** | 9.32E-01 | 4.70E-01 | 4.78E-02 | 5.14E-01 | **1.69E-10** | **3.90E-05** | 1.85E-01 | 3.95E-01 | 1.66E-01 |
|  | PC8 (~2.0%) | 1.00E+00 | **9.35E-48** | 3.27E-02 | 9.80E-01 | **1.01E-05** | **7.35E-04** | 7.12E-01 | **5.93E-33** | **2.05E-14** | **4.69E-04** | 7.52E-01 | 8.69E-02 |
|  | PC9 (~1.7%) | 1.00E+00 | **1.24E-08** | **4.57E-56** | 9.51E-01 | 8.67E-01 | 6.46E-01 | 9.07E-01 | **1.08E-07** | 5.96E-02 | 9.97E-01 | 9.41E-01 | 5.72E-01 |
|  | PC10 (~1.5%)­ | 1.00E+00 | **8.92E-43** | **4.24E-11** | 8.24E-01 | **8.49E-07** | 1.28E-01 | 6.08E-01 | **5.88E-14** | **1.25E-08** | **2.82E-03** | 1.56E-02 | **1.76E-03** |
| **Standardised +**  **Bimodal Signal Adjustment +  Filtering  (without IPS adjustment)** | PC1 (~49.0%) | 1.00E+00 | **2.58E-12** | 6.90E-02 | 8.07E-01 | **2.70E-02** | 9.54E-01 | 1.63E-01 | **5.82E-05** | **1.36E-05** | 1.03E-01 | 2.50E-01 | **2.14E-25** |
|  | PC2 (~5.0%) | 1.00E+00 | **6.02E-40** | **3.10E-02** | 9.97E-01 | **2.08E-04** | 3.18E-02 | 9.32E-02 | **7.22E-03** | **1.38E-02** | 7.05E-02 | **3.91E-09** | **5.09E-11** |
|  | PC3 (~3.6%) | 1.00E+00 | **1.51E-10** | 1.62E-01 | 8.91E-01 | **2.38E-03** | **1.71E-03** | 8.52E-01 | **8.93E-23** | **7.55E-09** | **2.13E-10** | **3.49E-11** | 2.63E-01 |
|  | PC4 (~2.6%) | 1.00E+00 | **8.41E-11** | **1.83E-08** | 9.94E-01 | **1.79E-06** | 8.79E-02 | 8.63E-02 | **5.07E-18** | **2.60E-09** | 8.80E-01 | **1.12E-05** | **1.59E-02** |
|  | PC5 (~2.1%) | 1.00E+00 | **1.62E-05** | 7.73E-01 | 9.57E-01 | **1.26E-03** | **1.38E-04** | **5.43E-05** | **2.54E-95** | **7.29E-20** | **2.80E-04** | 1.67E-01 | **2.89E-05** |
|  | PC6 (~1.6%) | 1.00E+00 | **1.30E-36** | **1.43E-02** | 9.34E-01 | 4.78E-02 | 9.04E-01 | 6.36E-01 | **4.11E-14** | **1.40E-08** | 3.78E-02 | **5.08E-08** | **2.05E-31** |
|  | PC7 (~1.5%) | 1.00E+00 | 6.89E-02 | **3.01E-04** | 9.34E-01 | 2.77E-01 | **4.37E-05** | **3.35E-03** | **2.58E-24** | **1.65E-09** | 2.13E-01 | **1.80E-02** | 1.07E-01 |
|  | PC8 (~1.3%) | 9.98E-01 | **1.79E-06** | **1.05E-46** | 8.78E-01 | 4.30E-01 | **4.64E-04** | 7.68E-02 | **1.62E-06** | **6.18E-05** | 2.22E-01 | 7.70E-02 | 6.66E-02 |
|  | PC9 (~1.1%) | 1.00E+00 | **5.03E-76** | **2.72E-02** | 9.09E-01 | **1.15E-04** | **3.52E-03** | 2.49E-01 | **1.71E-17** | **2.03E-10** | **3.87E-06** | 5.25E-02 | 1.11E-01 |
|  | PC10 (~0.9%) | 1.00E+00 | **4.21E-23** | **6.46E-34** | 9.54E-01 | 1.07E-01 | 4.42E-01 | 7.40E-01 | **3.54E-06** | **1.08E-02** | 2.86E-01 | 1.76E-01 | 9.79E-01 |
| **Standardised +**  **Bimodal Signal Adjustment +  IPS adjustment  (without Filtering)** | PC1 (~16%) | 1.00E+00 | **9.75E-13** | **2.96E-04** | 9.98E-01 | **3.10E-04** | 3.32E-01 | 1.27E-01 | **1.04E-03** | **8.20E-05** | **2.20E-04** | **6.21E-03** | **1.25E-10** |
|  | PC2 (~8.1%) | 1.00E+00 | **6.61E-07** | **1.79E-04** | 9.96E-01 | **2.28E-05** | 3.62E-01 | 9.32E-02 | **5.19E-12** | **3.75E-06** | 1.51E-01 | 5.40E-01 | 9.80E-01 |
|  | PC3 (~5.1%) | 1.00E+00 | **9.23E-25** | 1.73E-01 | 1.00E+00 | **3.22E-10** | **5.85E-08** | 1.48E-01 | **5.56E-67** | **2.00E-21** | **1.48E-07** | **4.60E-20** | **8.73E-08** |
|  | PC4 (~4.5%) | 1.00E+00 | **1.46E-19** | 9.43E-01 | 9.92E-01 | 5.10E-01 | 5.38E-01 | **1.27E-04** | **1.90E-28** | **3.22E-06** | 5.02E-02 | **2.94E-04** | **1.17E-03** |
|  | PC5 (~3.6%) | 1.00E+00 | **1.32E-08** | **4.06E-07** | 9.86E-01 | 9.56E-01 | 4.60E-02 | 4.13E-02 | **2.00E-08** | **8.61E-04** | 1.46E-01 | 6.44E-02 | **1.05E-13** |
|  | PC6 (~3.1%) | 1.00E+00 | **1.09E-06** | **5.32E-23** | 9.86E-01 | **3.50E-03** | **5.83E-06** | **2.28E-04** | **2.25E-09** | **4.53E-12** | 1.04E-01 | **5.67E-08** | **1.04E-11** |
|  | PC7 (~2.6%) | 1.00E+00 | **6.96E-06** | **4.54E-11** | 9.85E-01 | 1.35E-02 | 5.67E-01 | 6.90E-01 | **7.14E-21** | **5.27E-07** | 2.34E-01 | 1.16E-01 | **2.69E-06** |
|  | PC8 (~2.2%) | 1.00E+00 | **9.18E-56** | **5.48E-33** | 9.88E-01 | 1.46E-02 | **2.19E-03** | 2.41E-02 | **1.57E-14** | **1.62E-09** | **6.87E-04** | 2.93E-01 | **1.34E-04** |
|  | PC9 (~1.9%) | 1.00E+00 | 1.17E-01 | **4.79E-31** | 9.87E-01 | 4.82E-01 | 4.95E-02 | 3.22E-01 | **4.02E-11** | **7.81E-03** | 1.46E-01 | 9.02E-01 | 7.49E-01 |
|  | PC10 (~1.6%) | 1.00E+00 | **1.50E-29** | **1.63E-19** | 9.52E-01 | **3.43E-03** | 3.81E-01 | 8.57E-01 | **1.07E-05** | **7.05E-05** | 3.56E-02 | 1.22E-01 | **1.96E-06** |
| **Standardised +**  **Bimodal Signal Adjustment  (without IPS adjustment without filtering)** | PC1 (51.0~%) | 1.00E+00 | **1.22E-10** | 9.69E-02 | 1.00E+00 | 2.23E-01 | 8.66E-01 | 2.17E-01 | **4.93E-04** | **2.72E-04** | 1.61E-01 | 9.28E-01 | **1.80E-27** |
|  | PC2 (~4.6%) | 1.00E+00 | **6.07E-19** | 9.79E-02 | 9.99E-01 | **2.04E-05** | 5.21E-02 | 2.72E-02 | **1.40E-05** | **5.34E-03** | 5.98E-01 | **3.62E-05** | **3.71E-07** |
|  | PC3 (~3.6%) | 1.00E+00 | **1.93E-05** | **4.46E-06** | 9.40E-01 | **1.41E-02** | 1.80E-01 | 4.83E-01 | **3.55E-12** | **2.52E-07** | **1.79E-07** | 5.91E-01 | **3.36E-05** |
|  | PC4 (~3.2%) | 1.00E+00 | **2.88E-29** | **3.26E-05** | 1.00E+00 | **2.33E-07** | **1.42E-03** | 4.29E-01 | **3.19E-36** | **5.32E-12** | **5.56E-05** | **1.03E-17** | 3.80E-02 |
|  | PC5 (~2.6%) | 1.00E+00 | **2.42E-05** | 5.36E-01 | 9.67E-01 | **2.94E-04** | **2.80E-04** | **6.72E-05** | **1.84E-88** | **2.13E-21** | **1.13E-02** | 7.59E-01 | **2.84E-03** |
|  | PC6 (~1.5%) | 1.00E+00 | **1.43E-05** | **1.79E-02** | 9.82E-01 | **9.78E-03** | **1.72E-02** | 7.43E-01 | **1.68E-38** | **4.32E-14** | **1.75E-03** | 3.95E-01 | 1.18E-01 |
|  | PC7 (~1.4%) | 1.00E+00 | **2.73E-46** | 5.02E-02 | 9.73E-01 | 4.58E-02 | 5.44E-01 | 9.88E-01 | **1.88E-11** | **1.77E-05** | 4.01E-01 | **1.41E-08** | **1.57E-32** |
|  | PC8 (~1.4%) | 9.99E-01 | **6.34E-15** | **2.27E-147** | 9.34E-01 | 6.10E-01 | **7.36E-03** | 7.17E-01 | 9.03E-02 | **2.49E-02** | 5.81E-01 | 5.90E-01 | 9.55E-02 |
|  | PC9 (~1.2%) | 1.00E+00 | **2.18E-31** | **5.87E-08** | 9.53E-01 | 2.78E-02 | **5.06E-07** | **8.49E-06** | **1.36E-14** | **2.14E-09** | **3.73E-05** | **1.14E-04** | 1.05E-01 |
|  | PC10 (~0.9%) | 1.00E+00 | **2.16E-05** | **5.83E-08** | 9.96E-01 | **1.63E-02** | 4.68E-01 | 3.59E-01 | **6.26E-09** | **2.54E-03** | 2.25E-02 | **9.32E-03** | 6.95E-01 |
| **Standardised  Data** | PC1 (48%) | **8.71E-07** | **2.00E-09** | 3.41E-01 | **1.27E-02** | 4.84E-01 | 8.63E-01 | 2.15E-01 | **1.38E-03** | **7.53E-04** | 5.94E-02 | 9.22E-01 | **4.31E-29** |
|  | PC2 (7.2%) | 2.15E-01 | **4.92E-10** | 5.73E-01 | 8.88E-02 | **1.15E-07** | 2.61E-01 | 1.65E-01 | **<2.23e-308** | **1.95E-56** | **4.50E-12** | 9.37E-01 | 7.48E-02 |
|  | PC3 (4.5%) | **3.70E-03** | **1.41E-04** | 4.91E-02 | 2.90E-01 | 1.99E-01 | 4.35E-01 | 3.96E-02 | **3.80E-04** | 9.21E-02 | 1.31E-01 | 7.76E-02 | 9.18E-02 |
|  | PC4 (3.6%) | **4.35E-02** | **2.44E-13** | 1.27E-01 | 6.09E-02 | **1.88E-02** | 8.88E-02 | **1.78E-02** | **1.85E-12** | **3.72E-04** | 6.55E-01 | **6.54E-06** | **8.65E-16** |
|  | PC5 (2.9%) | **3.59E-09** | **3.13E-31** | 2.07E-01 | 8.85E-01 | **2.19E-03** | **1.62E-02** | **1.86E-02** | **1.82E-16** | **4.38E-06** | **2.71E-03** | **1.48E-14** | 3.15E-01 |
|  | PC6 (1.6%) | **7.62E-09** | **3.12E-08** | 2.35E-01 | 5.40E-02 | **2.63E-10** | **3.44E-04** | 5.45E-02 | **1.41E-34** | **1.54E-13** | 7.17E-01 | **6.94E-03** | **1.42E-05** |
|  | PC7 (1.3%) | **5.99E-09** | **1.48E-22** | **5.17E-04** | **2.69E-03** | **1.54E-02** | 8.47E-01 | 5.10E-01 | **2.15E-10** | **1.11E-04** | **1.51E-02** | **3.64E-06** | **2.63E-25** |
|  | PC8 (1.3%) | **1.28E-49** | **1.62E-04** | **5.71E-49** | **1.97E-06** | 4.71E-01 | **8.60E-06** | **6.14E-03** | **3.77E-06** | **6.37E-06** | 3.62E-02 | **8.15E-03** | **6.18E-03** |
|  | PC9 (1.1%) | **9.52E-05** | **2.91E-76** | **4.13E-12** | **1.96E-03** | **6.28E-03** | **1.47E-02** | **2.13E-05** | **1.86E-07** | **1.56E-06** | **1.08E-04** | 1.06E-01 | **3.08E-07** |
|  | PC10 (0.9%) | **7.52E-08** | **2.29E-09** | **6.19E-11** | **5.66E-05** | **1.41E-02** | 1.21E-01 | 6.18E-02 | **6.38E-10** | **1.43E-03** | 2.59E-01 | 3.54E-02 | 5.45E-01 |

**Table S7. *Associations between technical confounders and top 10 PCs.***The impact of technical confounders on each of the top 10 PCs of the two final sets of log abundance data (batch corrected, non-IPS adjusted and filtered, and batch corrected, IPS adjusted and filtered), measured using p-values from linear regression. In each case, the principal component was the dependent variable, and the technical variable was the independent variable, with some technical variables treated as linear predictors (number of freeze thaw cycles, sample age, sample volume, visual blood straining grade) and the rest treated as categorical variables. For technical variables that differed systematically by cohort (tranche number, previous freeze-thaw, number of freeze-thaw cycles, processing batch, treatment date, sample age, sample volume and blood staining), we included cohort as a covariate. Further details on the technical variables used assessed and their definitions are shown in Table S2. Entries with Benjamini-Hochberg adjusted p< 0.05 are bolded. Sample refers to samples of synovial fluid. Abbreviations: PC, principal component; IPS, intracellular protein score.
