## Supplementary Methods for "Methodological development of molecular endotype discovery from synovial fluid of individuals with knee osteoarthritis: the STEpUP OA Consortium"

**Consortium structure and governance**

Founding members of the STEpUP OA Consortium signed the Consortium Agreement in October 2019 and associated Data Sharing Agreement (see Consortium membership). An accession document allowed new joiners. The consortium was led by the University of Oxford (principal lead, Vincent), with samples and linked proteomic and clinical data being received, processed and held there. Proteomic analysis was by SomaLogic, Boulder, Colorado, US. Distinct working groups oversaw key activities according to pre-defined Terms of Reference (available on request) (Figure S1). Individuals analysing the data within the Data Analysis Group additionally signed a memorandum of understanding. A webpage included a lay summary and video (https://www.kennedy.ox.ac.uk/oacentre/stepup-oa/stepup-oa).

**Ethics and cohorts**

University of Oxford Medical Sciences Central University Research Ethics Committee (CUREC) granted ethical approval for the processing, storage and use of samples and linked data for this project on 1^st^ November 2019 (R67029/RE001). Each site ensured compliance with local ethical and data protection policies and appropriate written informed consent from each participant via existing approvals which included collaborative use. All signed a material transfer agreement prior to movement of samples/data.

To ensure confidentiality and to generate a single dataset, secondary pseudo-anonymisation was performed by the Oxford site for all received samples and their associated data records, with secure records of linkage with the originating cohort’s participant identification numbers. Specifically, a STEpUP participant ID number (PIN) and related unique sample identification number (SIN) were generated for each participant and associated sample(s), which did not identify cohort.

**Laboratory methods**

Tranches were processed as samples were received, Tranche 1-4 sample numbers were 435, 610, 691, and 10 samples respectively. Samples and enzyme were kept in pre-cooled blocks once thawed until use. ‘Piston’ pipettes (Gilson, UK) were used to ensure accurate measurement of synovial fluid.

**Clinical data**

Radiographic severity was defined by ordinal Kellgren and Lawrence (KL) grading for the worst affected compartment (0-4), as provided by the cohort. X-rays were not transferred or re-scored. Where individual KL grades were not available, dichotomous variables were generated, based on each cohort’s eligibility criteria, i.e. stating presence or absence of radiographic knee OA (KL≥2) and/or presence or absence of advanced radiographic knee OA (KL scores KL≥3) (Table S2).

The Western Ontario and McMaster Universities Osteoarthritis Index (WOMAC)^[1]^, the Knee Injury and Osteoarthritis Outcome Score (KOOS)^[2]^ and various knee pain numerical rating scales were most commonly collected by cohorts. No single patient reported outcome measure was held by all participating cohorts. In a systematic review and meta-analysis carried out by consortium investigators specifically to support the harmonisation of its knee pain data^[3]^, the ability to define a harmonised knee pain threshold by analysis of Patient Acceptable Symptom State (PASS) for these subscales in these populations was explored^[3]^. These findings supported the generation of a dichotomous harmonised pain category for the purposes of the consortium work (those with unacceptable or acceptable levels of knee pain, based on a single threshold). Pain scale selection from available cohort data followed an algorithm which prioritised WOMAC pain subscale for OA cases and KOOS pain subscale for knee injury cases, followed by knee pain NRS, followed by painDETECT NRS^[4]^ where others were unavailable. These same harmonizable knee pain outcomes were also used continuously where available.
