## Supplementary Figures for "Methodological development of molecular endotype discovery from synovial fluid of individuals with knee osteoarthritis: the STEpUP OA Consortium"

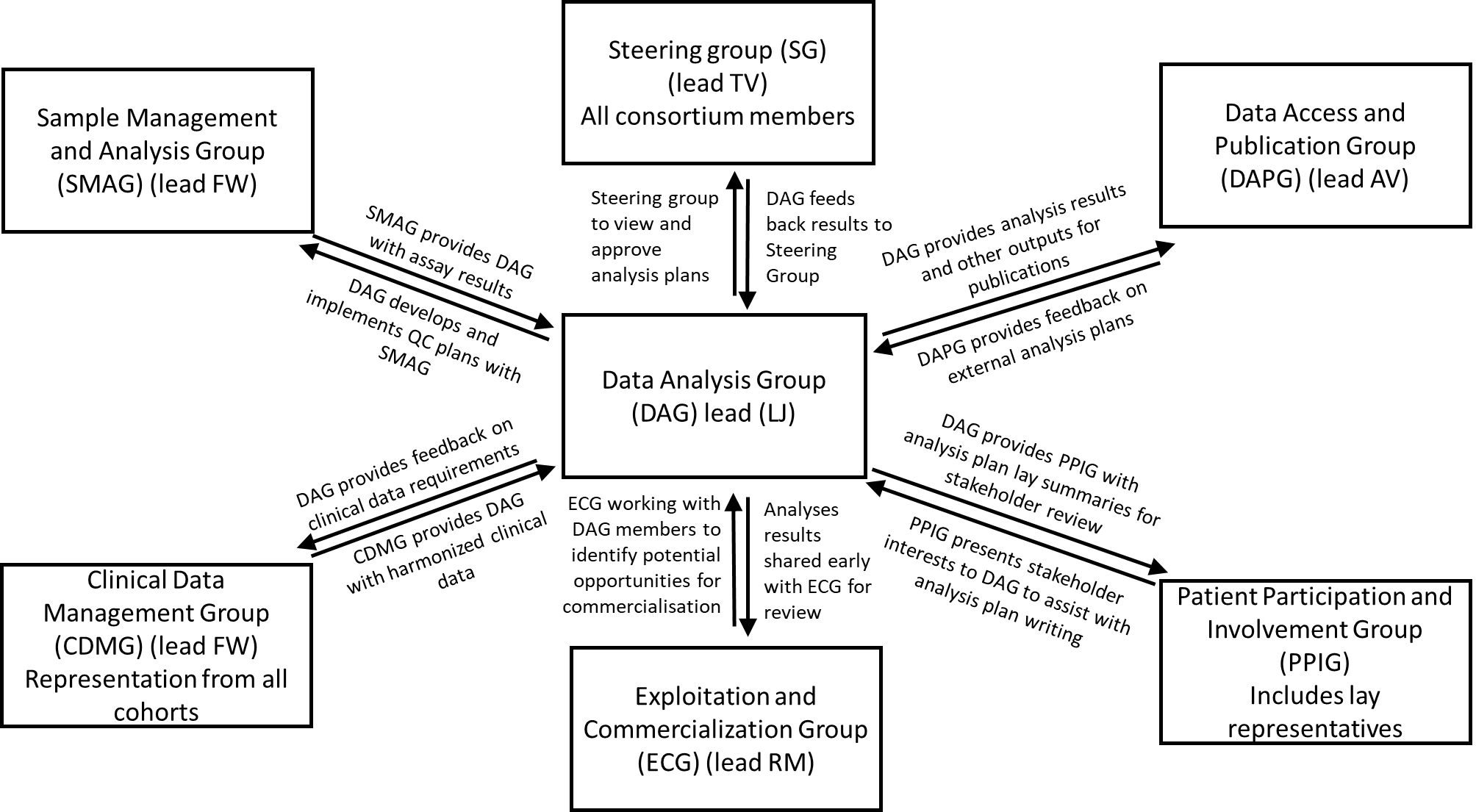

**Figure S1.** Consortium structure, as working groups. Distinct working groups oversaw key activities according to pre-defined Terms of Reference (available on request). TV, Tonia Vincent; FW, Fiona Watt; AV, Ana Valdes; LJD, Luke Jostins-Dean; RM, Rose Maciewicz.

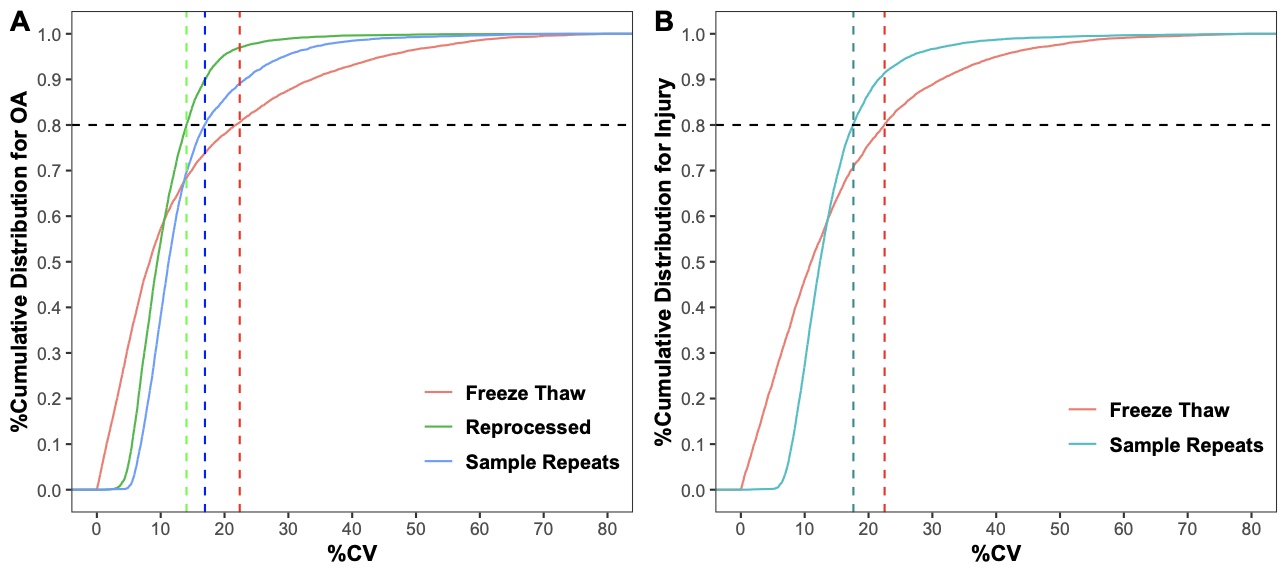

**Figure S2.** Assessment of assay repeatability using pooled samples of synovial fluid from participants with (A) knee OA and (B) acute knee injury, measured by the coefficient of variation (%CV). These include the repeatability of the standard processed pooled samples included on every plate (‘Sample Repeats’), pooled samples which had been repeatedly freeze-thawed (‘Freeze Thaw’) prior to processing and an OA pool aliquot that had been freshly enzyme digested with stored hyaluronidase during each of the 2nd, 3rd and 4th tranches of sample processing (done for the OA pool only) (‘Reprocessed’). Dotted vertical lines show the maximum %CV for 80% of proteins for each group.

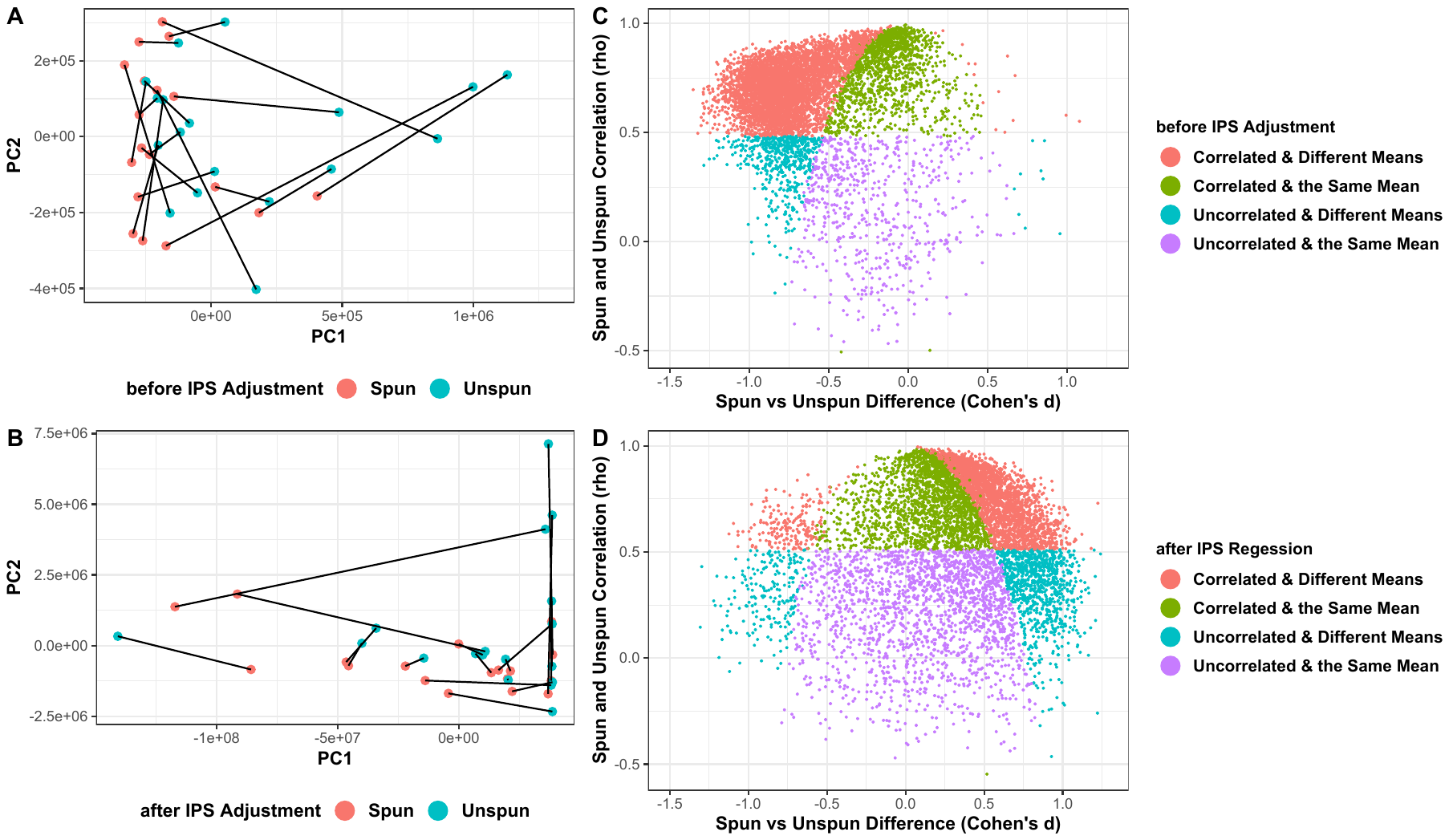

**Figure S3.** Top 2 principal components of (A) non-IPS adjusted and (B) IPS adjusted log RFU of the 18 pairs of centrifuged (spun) and non-centrifuged (unspun) SF samples. Samples are coloured by spin status and paired samples are linked by lines. Measures of differential abundance (Cohen's d) and Pearson correlation coefficient (rho) between spun and unspun samples for (C) non-IPS adjusted and (D) IPS adjusted log RFU. Samples are coloured depending on their significance (Benjamini-Hochberg adjusted p< 0.05) on the two measures: Different Means corresponds to a significant difference in means in a paired t-test and Correlated corresponds to a significant correlation in a Pearson correlation test. IPS, intracellular protein score; PC, principal component; SF, synovial fluid; RFU, relative fluorescence unit.

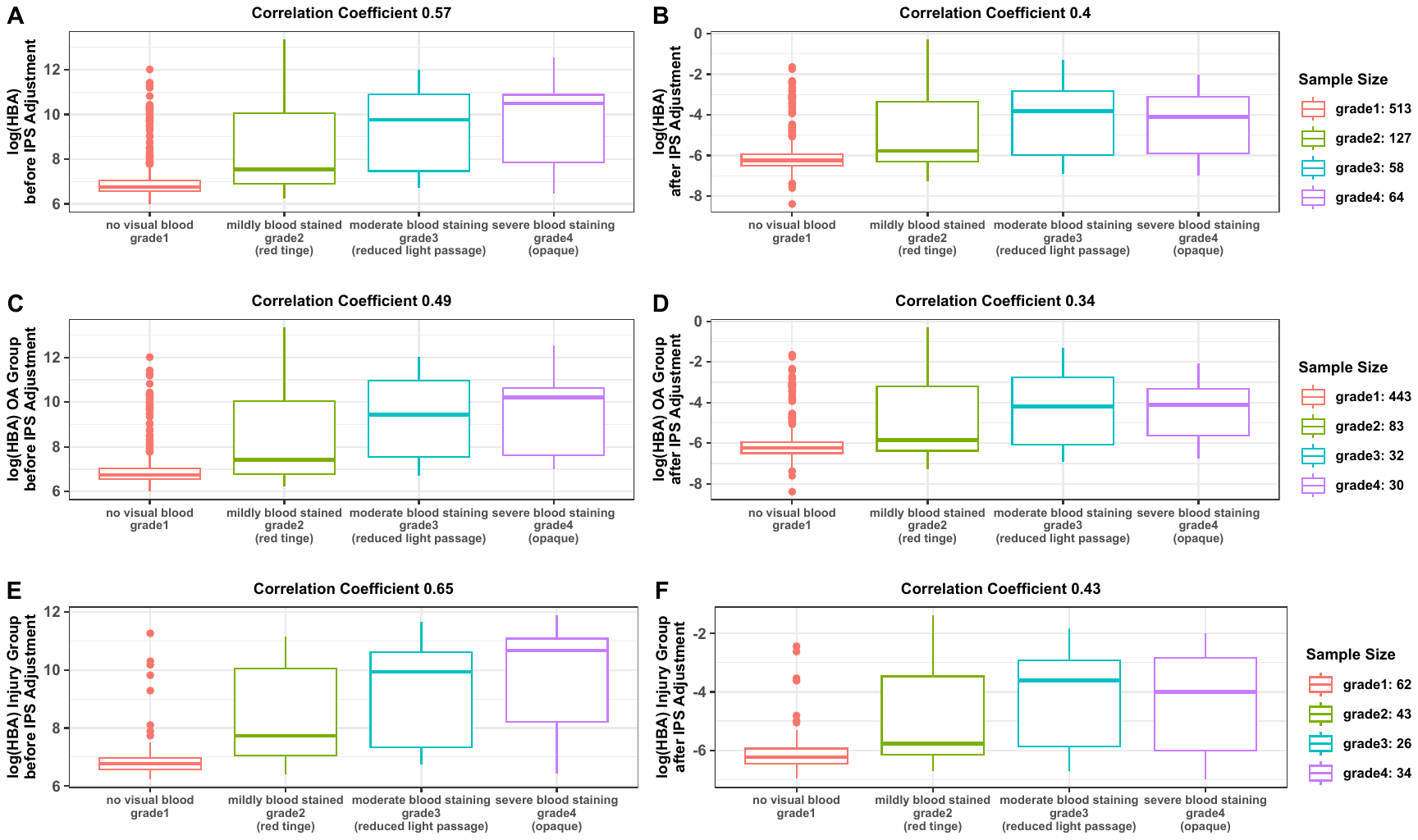

**Figure S4**. Boxplots showing the correlation between visual blood staining grade of SF at the time of sample collection and the blood analyte, HBA, in non-IPS adjusted data in (A) all samples, (C) OA samples and (E) acute knee injury samples, and in IPS adjusted data in (B) all samples, (D) OA samples and (F) acute knee injury samples. Spearman correlation coefficients measuring rank-based correlation considering visual blood staining as an ordinal variable are shown. 443 OA samples had blood staining grade 1 (no blood detected, 75% among the 588 total samples with blood staining records). HBA, haemoglobin A; IPS, intracellular protein score; SF, synovial fluid.

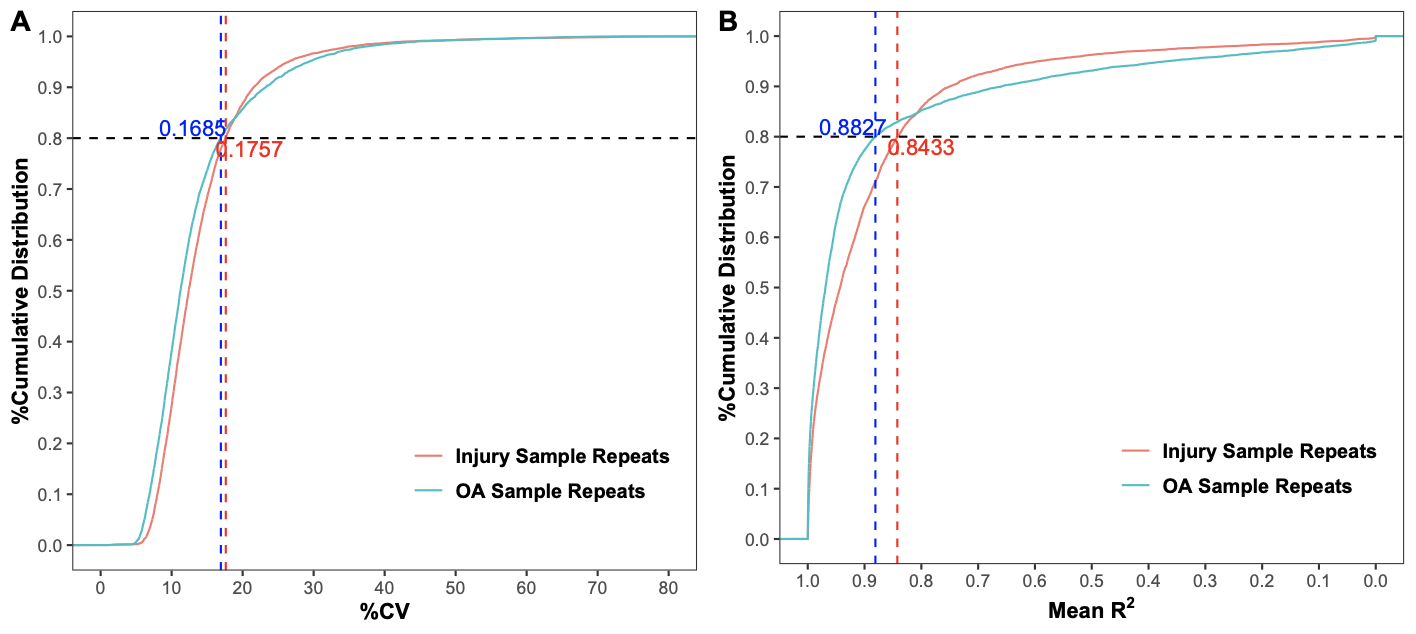

**Figure S5.** (A) Assessment of assay repeatability after optimised quality control procedures measured using the cumulative distribution of the coefficient of variation (%CV) on pooled OA samples (OA Sample Repeats) and pooled acute knee Injury samples (Injury Sample Repeats) separately. 80% of proteins had a %CV less than 16.85% and 17.57% in the OA and acute knee injury pools (blue and red dotted lines respectively). (B) The proportion of variation that was estimated to be non-technical, measured by R^2^ for OA and acute knee injury sample repeats separately. 80% of proteins had R^2^ values greater than 88.27% and 84.33% in the OA and knee injury pools (blue and red dotted lines respectively).

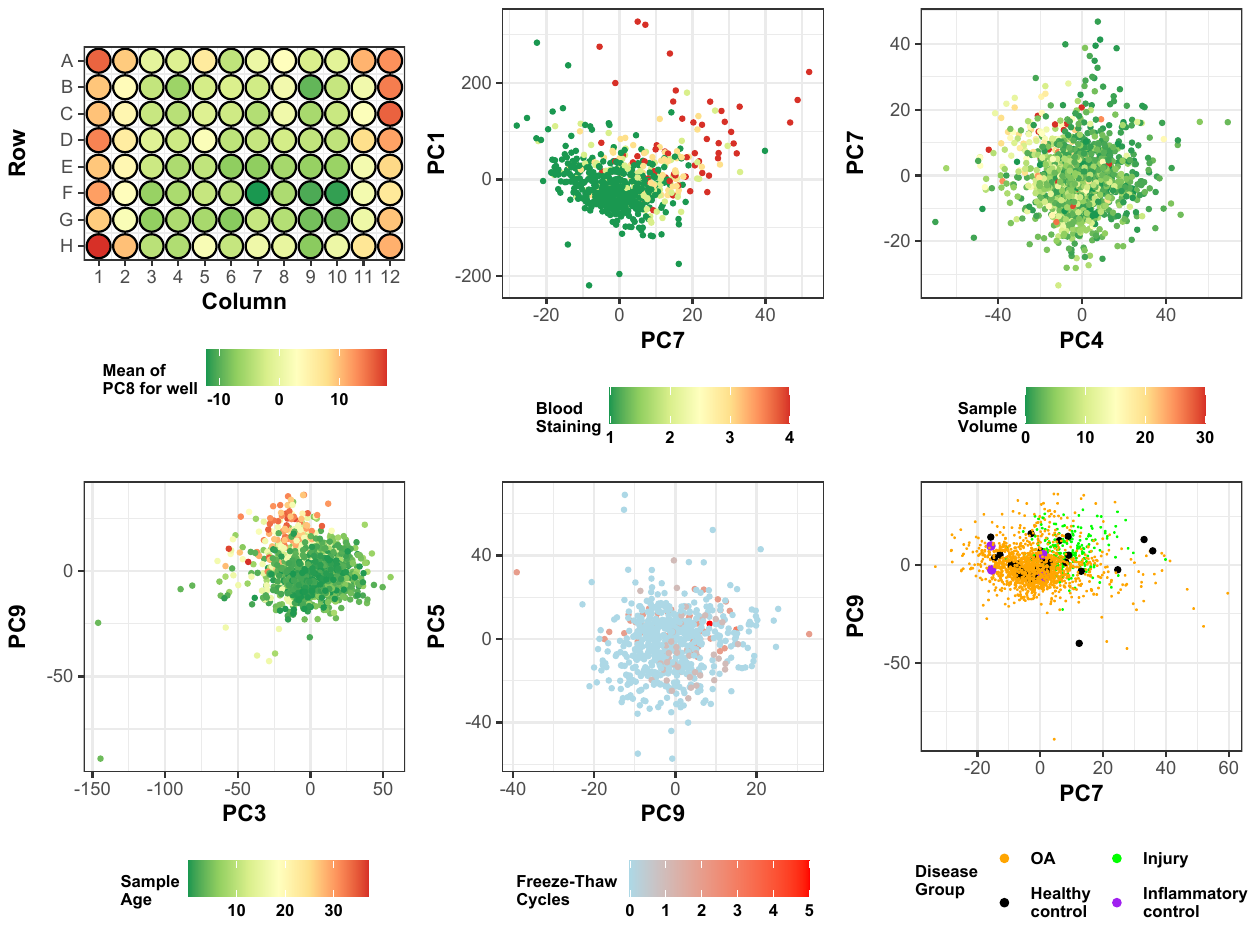

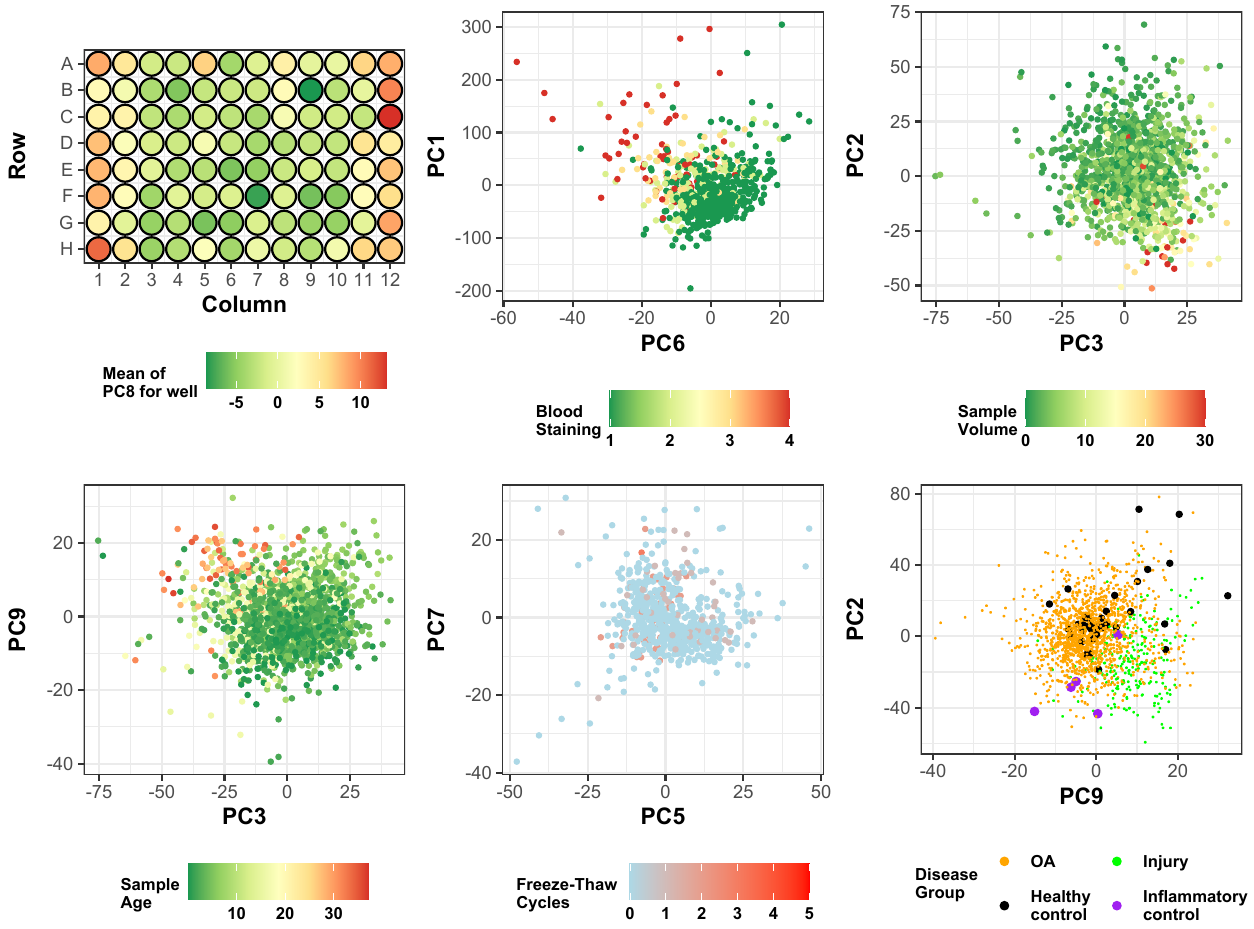

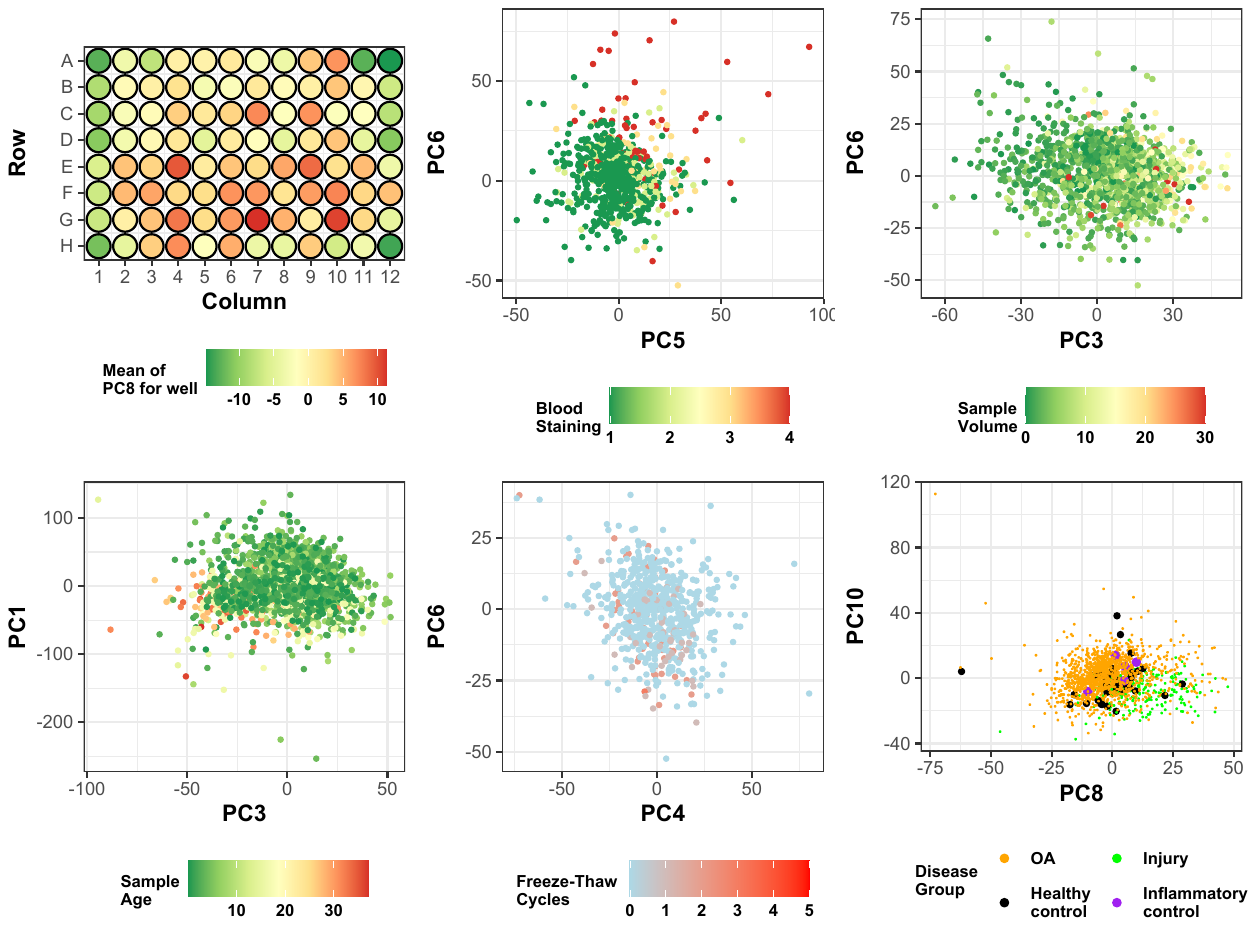

**A**

**B**

**C**

**Figure S6.** Visualisation of pre-defined technical confounders by select principal components of the (A) non-filtered IPS adjusted (B) filtered non-IPS adjusted and (C) non-filtered non-IPS adjusted data. Visualisation of the two PCs most strongly associated with each confounder (colours correspond to confounder value). Confounders include plate position (mean of PC8), blood staining grade of sample (which was performed immediately after aspiration from the joint by visual inspection), volume of sample taken during aspiration, age of the sample in years from aspiration to processing, the number of times the sample had been thawed and re-frozen, the disease group of the sample (knee OA, acute knee injury, healthy control, inflammatory arthritis control). The association between each PC and confounder is shown in the Table S7.

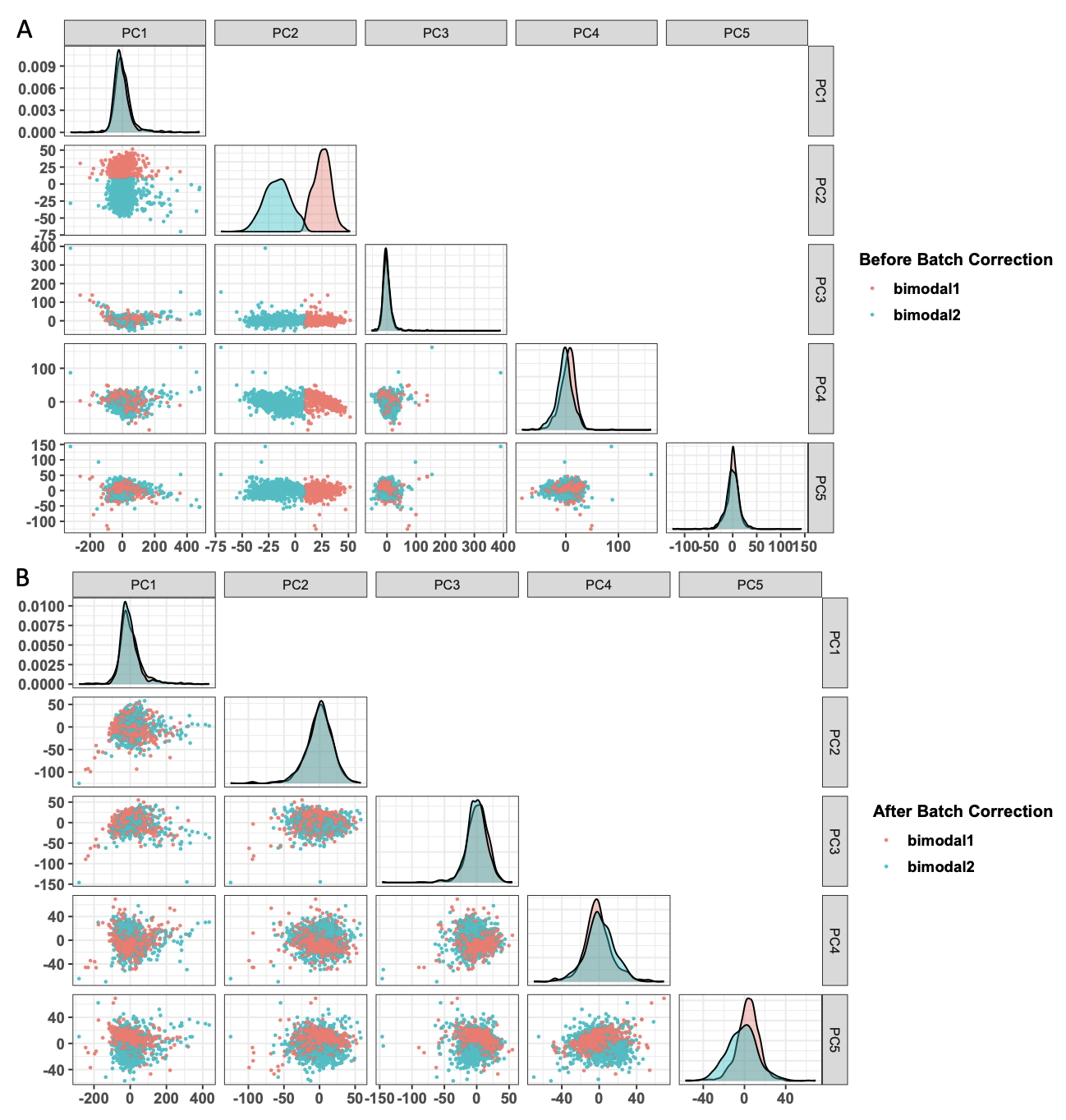

**Figure S7.** Pairwise scatter plots (off-diagonal) and histograms (diagonal) of the top five principal components of standardised log abundance, (A) before and (B) after batch correction for plate and bimodal signal status, coloured by bimodal signal status. Batch correction effectively removed the effect of bimodal signal status on the top PCs.
